# Endoleak Prediction After EVAR: A Point Cloud Neural Network Framework Enhanced by Computational Fluid Dynamics and Multi-Features

**DOI:** 10.64898/2026.01.27.26345009

**Authors:** Chen Peng, Yaoyi Zhang, Weifeng Guo, Lingwei Zou, Zhihui Dong, Jun Jiang, Wei He

**Affiliations:** Department of Cardiology, The Second Affiliated Hospital, School of Medicine, Zhejiang University, Hangzhou, 310009, China; State Key Laboratory of Transvascular Implantation Devices, 310009, Hangzhou, China; Heart Regeneration and Repair Key Laboratory of Zhejiang Province, Hangzhou, 310009, China; Transvascular Implantation Devices Research Institute, Hangzhou, 310053, China; Department of Vascular Surgery, Zhongshan Hospital, Fudan University, Shanghai, China; Institute of Vascular Surgery, Fudan University, Shanghai, China; National Clinical Research Center for Interventional Medicine, Zhongshan Hospital, Fudan University, Shanghai, China; Department of Radiology, Zhongshan Hospital, Fudan University, Shanghai, China; Department of Cardiology, Zhongshan Hospital, Fudan University, Shanghai Institute of Cardiovascular Diseases, China; State Key Laboratory of Cardiovascular Diseases, Zhongshan Hospital, Fudan University; NHC Key Laboratory of Ischemic Heart Diseases

**Keywords:** Point Cloud Neural Network, Endoleak, Abdominal Aortic Aneurysm, Multimodal Features, Computational Fluid Dynamics

## Abstract

**Background:** Endovascular aortic aneurysm repair (EVAR) is effective in preventing rupture of abdominal aortic aneurysm (AAA), but endoleak remains a serious postoperative complication. Accurate prediction of endoleak risk is a significant clinical challenge.

**Purpose:** This study aimed to evaluate the value of a Point Cloud Neural Network (PCNN) in predicting endoleaks after EVAR by integrating multimodal features.

**Materials and Methods:** We collected follow-up data from 381 AAA patients. Radiomic characteristics of the procedural intraluminal thrombus and morphological parameters were extracted following medical image segmentation and 3D reconstruction. Hemodynamic parameters, including time-averaged wall shear stress (TAWSS), oscillatory shear index (OSI), and relative residence time (RRT), were obtained through a semi-automated computational fluid dynamics (CFD) workflow.

Six traditional machine learning models and four PCNN architectures were developed with progressively added feature sets: 1) medical history and morphology (H+M); 2) H+M+R; 3) H+M+CFD; and 4) all features combined (H+M+R+CFD).

**Results:** Traditional ML models showed limited performance (AUC range: 0.55-0.77). In contrast, PCNN models demonstrated substantially improved predictive capability. The baseline PCNN (H+M) achieved an AUC of 0.81. The RA-PCNN model incorporating radiomic features showed a 6.58% improvement (AUC=0.86). The CFD-PCNN model with hemodynamic parameters exhibited a 13.0% increase (AUC=0.91), with superior F1-score (0.78) and recall (0.88). The multimodal RA-CFD-PCNN model performed best, achieving an AUC of 0.93, accuracy of 0.90, and F1-score of 0.83.

**Conclusion:** This study establishes a PCNN-based framework for endoleak prediction that significantly outperforms traditional machine learning methods, providing an effective approach for assessing endoleaks in AAA patients.

**Summary statement:** This study developed a PCNN-based framework integrating clinical, morphologic al, radiomic, and hemodynamic features from 381 AAA patients to predict endoleaks after EVAR. Results demonstrated superior performance over traditional ML, with hemodynamic parameters providing a major performance boost, highlighting the value of physiological and biomechanical feature integration for vascular disease prediction.

**Key Results:** The multimodal PCNN model integrating all features achieved an AUC of 0.93, significantly outperforming traditional machine learning models (AUCs 0.55-0.77).

Incorporating hemodynamic parameters provided the greatest performance increase, with the CFD-PCNN model’s AUC increasing by 13.0% to 0.91 compared to the baseline PCNN (AUC=0.81).

The model combining radiomics and hemodynamics (RA-CFD-PCNN) achieved the highest F1-score of 0.83 and AUC of 0.93, demonstrating robust predictive accuracy.

## 1 Introduction

Abdominal aortic aneurysm (AAA) involves irreversible dilation of the abdominal aorta and above a certain threshold carries an associated high risk of rupture if untreated^[1]^. Current guidelines recommend intervention when the maximum diameter exceeds 55 mm^[2]^. Endovascular aortic aneurysm repair (EVAR) is a common treatment to prevent aneurysm growth and rupture; however, it can be complicated by endoleak—persistent blood flow into the aneurysm sac ^[3]^. As a major post-EVAR complication, endoleak increases the risk of continued expansion and rupture, often requiring secondary interventions^[4]^. Accurate prediction of endoleak can support the appropriate level of clinical evaluation. This recognizes the potential need for increased decreased surveillance post EVAR.

Patients undergoing EVAR require lifelong imaging surveillance, typically using Computed Tomography Angiography (CTA) as the gold standard. However, CTA monitoring carries risks of renal injury and radiation exposure^[5]^. While patient history provides broader clinical context, current guidelines rely heavily on oversimplified anatomical metrics like maximum diameter—an approach with documented limitations. A large cohort study by Varkevisser et al., found that 22.3% of patients meeting intervention criteria (diameter >5.5 cm) showed no expansion over 5 years, while 13.6% of subthreshold cases ruptured unexpectedly^[6]^. These discrepancies highlight the need to incorporate additional pathophysiological information beyond conventional morphological parameters to improve postoperative risk stratification.

Numerous studies have employed computational mechanics and numerical simulations to evaluate endoleak risk after EVAR by analyzing interactions between vascular dynamics, vessel walls, and blood flow. These approaches typically rely on transient simulations of biomechanical parameters to assess potential complications^[7,8]^. Helo et al. found that both abnormally high and low wall shear stress (WSS) can increase endoleak risk—low WSS may cause thrombosis, while high WSS can lead to stent displacement and reduced stability^[9]^ The oscillatory shear index (OSI) quantifies directional changes in wall shear stress. Elevated OSI values reflect unstable flow patterns, which may compromise arterial wall integrity and stent adhesion, thereby increasing endoleak risk^[10]^. It should be noted that the assessment of WSS and related parameters. While informative in research, is not routinely performed in standard clinical follow-up. Current protocols prioritize anatomical evaluation via imaging. While informative in research, is not routinely performed in standard clinical follow-up. Current protocols prioritize anatomical evaluation via imaging. The integration of these quantitative hemodynamic parameters into routine clinical decision-making awaits further technological simplification, standardization of acquisition protocols, and robust validation of their prognostic utility in large-scale prospective studies.

David et al. and Li et al. applied fluid-structure interaction models to examine interactions between blood flow, vascular walls, and stent grafts. By analyzing stress distributions and drag forces, they investigated stent migration mechanisms and potential factors contributing to endoleaks^[11–13]^. Zhang and Laubrie used constrained mixture theory to model vascular growth and remodeling after stent implantation. Their fluid-solid interaction approach quantitatively analyzed elastic protein degradation and collagen deposition in an idealized vascular model, providing insights into long-term endoleak mechanisms, though it did not account for complex blood flow patterns or realistic aneurysm anatomy^[14,15]^. The morphology and volume of the intimal thrombus can affect the hemodynamic environment of the collateral vessels, thereby affecting the risk of endoleaks^[8]^. These mechanistic models face several limitations, including high computational complexity, discrepancies from real physiological conditions, difficulties in accurately defining branch artery boundary conditions, and the substantial clinical resources required to obtain precise material parameters for the abdominal aorta^[16]^. However, the computational intensity of solving partial differential equations makes these models unsuitable for clinical timelines, while small sample sizes further limit their generalizability^[17,18]^.

Several studies have employed artificial intelligence (AI) algorithms combined with multimodal clinical data to develop less resource-intensive approaches for endoleak management compared to traditional mechanistic models. For example, Talebi et al. proposed an innovative method incorporating synthetic generation and removal of endoleak regions in CT images for data augmentation, integrated with a U-Net Convolutional Neural Network (CNN) architecture. Clinical validation demonstrated 95% diagnostic accuracy, significantly surpassing conventional methods while maintaining strong generalizability across diverse datasets^[19]^. Yang et al. developed a deep learning (DL) model for endoleak detection using non-contrast CT scans from 167 patients (85 with endoleaks, 82 without). The study found significantly higher Hounsfield unit (HU) values and greater density heterogeneity in the aneurysm sac of endoleak patients. The model achieved high sensitivity through automated segmentation and quantitative analysis of these imaging biomarkers^[20]^. Current AI-assisted detection methods have improved the speed and accuracy of postoperative endoleak identification, but they remain unable to predict endoleak risk prior to surgery. A preoperative risk prediction model is still needed to enhance long-term clinical management of AAA patients^[21]^. The construction of integrated diagnostic models based on multimodal data fusion and AI technology for predicting the onset and progression of cardiovascular diseases has become a research hotspot^[22]^. Multimodal data along with physiological, morphological, and biomechanical parameters provide complementary information that enables a more comprehensive understanding of pathophysiology. Integrating these data can enhance the predictive power of AI models and improve the diagnosis and risk stratification of cardiovascular diseases. For instance, Wang et al. introduced a two-stage multimodal fusion paradigm for cardiovascular disease diagnosis and screening, which demonstrated strong performance in both internal validation and external multicenter evaluations^[23]^. Pezel et al. developed a ML model integrating coronary CTA and stress cardiac Magnetic Resonance Imaging (MRI) data, which achieved an AUC of 0.86 in predicting major adverse cardiovascular events (MACE) in patients with obstructive coronary artery disease, outperforming conventional risk scores^[24]^. Kim et al. developed a DL framework that integrates multi-physical features—including biomechanical parameters, imaging data, and hemodynamic simulations—to predict abdominal aortic aneurysm progression. This approach enhances personalized preoperative risk assessment of AAA growth^[25]^. However, these AI-driven advances in cardiovascular diagnostics have primarily focused on preoperative assessment and have not yet been extended to the management of postoperative complications, particularly for endoleak management following EVAR. The proposed multi-modal AI framework is inherently extensible to preoperative data integration for the endoleak prediction. Specifically, the model architecture and feature-engineering pipeline are designed to accommodate multi features, including imaging radiomics, evolving morphological changes of the aneurysm sac, spatial hemodynamic alterations, and other clinical indicators. The pathogenesis of endoleaks manifests multidimensional correlations, encompassing not only local anatomical metrics (e.g., neck diameter, intraluminal thrombus (ILT) volume) but also systemic factors including patient-specific comorbidities, hemodynamic distributions, and biomarker trajectories. This pathophysiological complexity necessitates multimodal data integration, where computational synthesis of cross-domain features enables precision risk stratification. Current research gaps persist in developing unified AI architectures capable of hierarchically fusing heterogeneous data streams, particularly for real-time endoleak prediction after EVAR surgery.

In this study, we employed ten AI methods to predict post-EVAR endoleaks in 381 AAA patients. Pre-operative features included medical history, automatically segmented morphological measures, and Radiomic features. We built six ML models. Support Vector Machine (SVM), eXtreme Gradient Boosting (XGBoost) and Logistic Regression, each in two versions: one using medical history and morphology, and another adding Radiomic (R). Additionally, a semi-automated CFD workflow computed hemodynamic parameters, and four DL point cloud networks were developed:(i) a point cloud neural network (PCNN) incorporating patient medical history(Stage 1: H+M), (ii) an RA-PCNN model that adds Radiomic features to the previous model(Stage 2: H+M+R), (iii) a CFD-PCNN (Stage 3: H+M+CFD) model incorporating some WSS-based hemodynamic parameters, and (iv) an RA-CFD-PCNN (Stage 4: H+M+R+CFD) model that integrates cross-modal features from all dimensions. Model performance was evaluated using AUC-receiver operating characteristic curve (ROC), precision-recall curves, and rank tests. A dual-axis analytical framework was applied: horizontal analysis compared ML versus neural network architectures with/without Radiomic, while vertical analysis assessed performance across four stages of feature integration (Stage 1 to Stage 4). By comparing these ten models, this work provides clinicians with a comprehensive reference for pre-operative risk assessment and personalized surgical planning for AAA patients.

## Materials and Methods

The overall process was shown on **Fig.1**. Briefly, Figure 1(a) illustrates CTA preprocessing and the 3D U-Net–based multi-class segmentation used to delineate the aneurysm lumen and ILT. Figure 1(b) shows ILT radiomics extraction using PyRadiomics, while Figure 1(c) presents the semi-automated extraction of anatomical features, including local morphologic measurements (e.g., diameters, angulation, tortuosity) and global shape descriptors derived from the statistical shape model. Figure 1(d) outlines the semi-automated CFD pipeline used to compute hemodynamic indices (TAWSS, OSI, and RRT) and derive spatially averaged parameters. Finally, Figure 1(e) summarizes the two predictive strategies, in which we trained (1) traditional ML baselines using tabular features and (2) PCNN-based models with progressive multimodal fusion (H+M, H+M+R, H+M+CFD, and H+M+R+CFD), followed by evaluation on a held-out test set.

**Figure 1.**
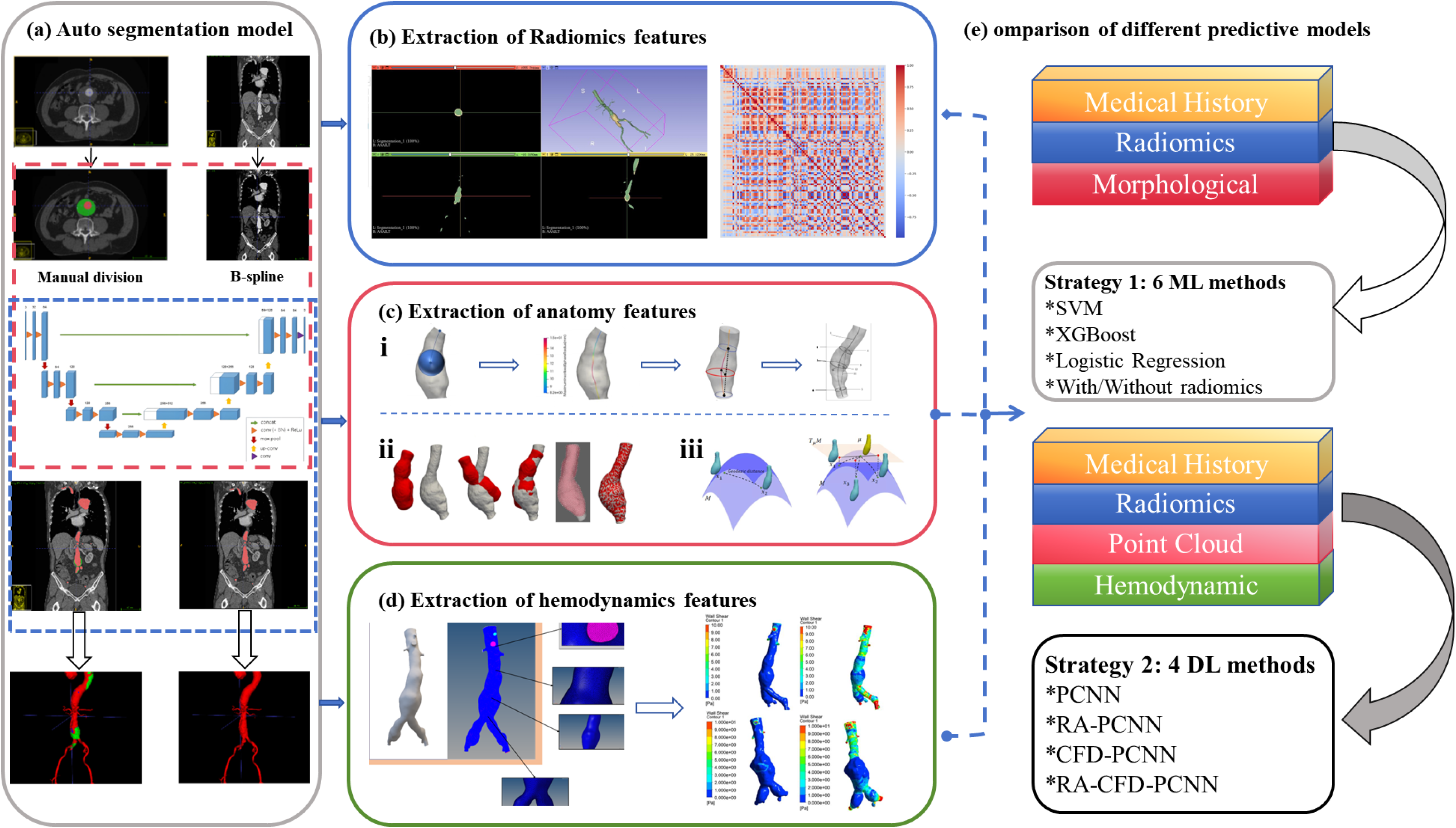
(a) 3D Unet training model. (b) Radiomics features extraction pipeline. (c) Anatomy features extraction pipeline. (d) Hemodynamics features extraction pipeline. (e) Two endoleak prediction strategy.

### Baseline Characteristics

A retrospective study was conducted from January 2016 to December 2019 at the Department of Vascular Surgery, Zhongshan Hospital, Fudan University. The study included patients diagnosed with infrarenal AAA by CTA or digital subtraction angiography (DSA), who underwent standard EVAR. The study protocol was approved by the Institutional Review Board of our hospital. Due to the retrospective nature of the investigation, the requirement for informed consent was waived. Strict confidentiality measures were implemented to protect patient privacy.

Inclusion and Exclusion Criteria:

(1) Inclusion criteria

a. Age ≥18 years
b. Diagnosis of infrarenal AAA confirmed by CTA or DSA
c. Meeting standard indications for EVAR with surgical consent, and undergoing the procedure between January 2016 and December 2019
d. Completion of standardized thoracoabdominal CTA scans preoperatively, and at 3/6/12 months postoperatively with annual follow-up thereafter
(2) Exclusion criteria

a. Prior aortic surgery history
b. Thoracoabdominal aortic aneurysm
c. Infectious AAA
d. Ruptured AAA
e. AAA with Marfan syndrome
f. Patients receiving chimney EVAR or fenestrated EVAR
g. Incomplete surgical records
h. Follow-up duration <1 year
i. Development of non-type I/II endoleaks during follow-up
j. Poor-quality imaging data precluding evaluation

Final cohort of 502 AAA patients initially screened, 381 met the eligibility criteria and were included in the analysis. The patient selection process is illustrated in **Fig. 2**.

**Figure 2.**
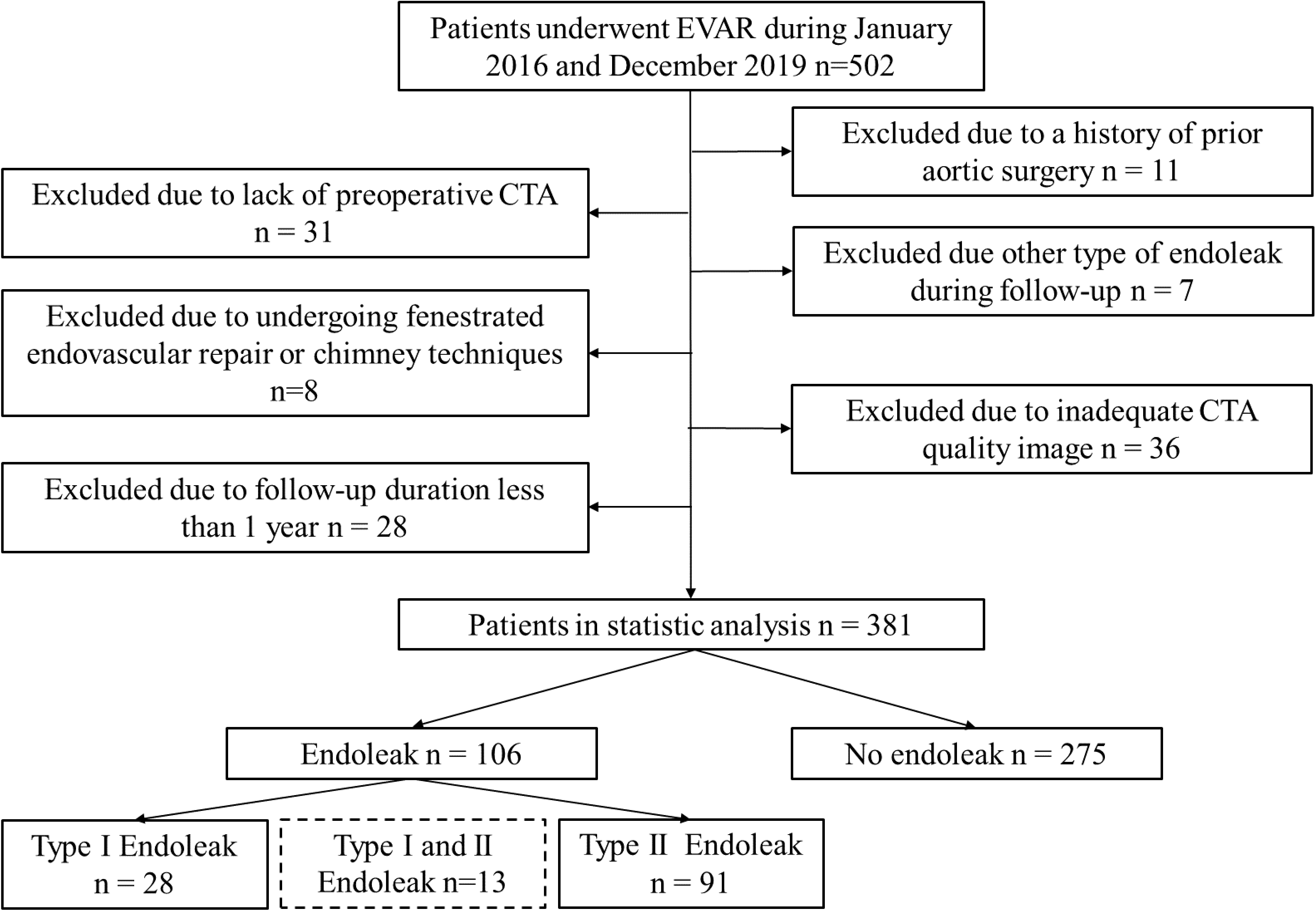
Patient inclusion and exclusion flowchart. The study cohort comprised 381 AAA patients (327 males, 54 females) with a mean age of 70±9 years, followed for 1,129±665 days. Patients were stratified into an endoleak group (n=106, 27.8%) and non-endoleak group (n=275, 72.2%), with the endoleak subgroup demonstrating 91 type II endoleaks, 28 type I endoleaks, and 13 cases exhibiting concurrent type I and II endoleaks.

### 3D U-Net Based Multi-Classification Automated Segmentation System for AAA

The complex 3D anatomy of AAA is critical for surgical risk assessment, yet existing medical imaging platforms (e.g., 3D Slicer, Mimics) often involve cumbersome workflows and limited automation. To address this, we developed an automated three-class segmentation system based on 3D U-Net, which accurately delineates the AAA lumen, wall thrombus, and background. This approach reduces reliance on extensive manual annotation and avoids complex preprocessing, enabling efficient end-to-end training.

Original DICOM images (512×512 pixels, 1.12 mm³ isotropic voxel size) underwent a two-stage preprocessing pipeline to enhance image fidelity and optimize feature representation for subsequent analysis. First, to mitigate partial volume effects and facilitate precise morphological delineation of the aortic wall and potential endoleaks, images were up sampled to a 1024×1024 matrix (0.56 mm ³ voxel size) using bicubic interpolation. This process improves spatial continuity and provides a higher-resolution matrix that is crucial for detailed vascular reconstruction and accurate biomechanical simulations. Subsequently, to accentuate clinically relevant boundaries and improve the model’s sensitivity to subtle contrast variations indicative of endoleaks, an unsharp mask filter was applied for edge enhancement. This step selectively increases the contrast at tissue interfaces and within lesion regions, thereby augmenting the discriminative information available. After that, manual segmentation was performed in ITK-SNAP by a vascular surgeon (≥5 years of experience) and verified by a senior expert (≥10 years of experience, the lead vascular radiologist on the study, with specialized expertise in EVAR follow-up imaging). The annotated region extended from the ascending aorta to the iliac bifurcation, including major branching vessels: brachiocephalic artery (BCA), left common carotid (LCCA), left subclavian (LSA), celiac trunk (CA), renal arteries (RRA/LRA), superior mesenteric artery (SMA), and iliac arteries (RIIA/REIA/LIIA/LEIA). Distinct labels were assigned to the contrast-enhanced lumen (red) and wall thrombus (green) for model training. The 3D U-Net architecture was employed for segmentation, the encoder (contraction path) consists of two 3×3 convolutional layers with batch normalization, Rectified Linear Unit (ReLU) activation, and max pooling. The decoder (expansion path) uses up-sampling layers followed by two 3×3 convolutions and ReLU to reconstruct image details. The model was trained using annotated vascular CT labels and original DICOM data, yielding a robust segmentation network. The final segmented region—containing the aneurysm and thrombus—was exported as a 3D geometric model.

The model training is performed using a multimodal loss function, combined with Dice coefficient loss function:

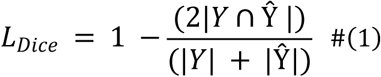

and weighted cross-entropy loss function:

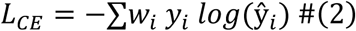

For joint optimization, category weights were dynamically adjusted based on the pixel proportion of vessel/aneurysm regions. The model was trained on a clinical peripheral vascular CT angiography dataset with expert-annotated labels for aneurysm and thrombus regions.

The overall 3D Unet work model for automatic segmentation for aneurysm and thrombus was shown in **Fig.1 (a).**

This automated segmentation provided the basis for subsequent extraction of radiomic and morphological parameters, as well as hemodynamic simulations.

### Extraction of Radiomic features

Previous studies indicate that the ILT in AAA plays a significant biomechanical role^[26–28]^, reducing wall stress and potentially preventing rupture^[29,30]^. Given the correlation between thrombus biomechanics and postoperative EVAR complications, we hypothesize that Radiomic features of the thrombus may predict endoleak.

In this study, Radiomic features of the thrombus were extracted using the PyRadiomics package in Python 3.7.16^[31]^ **(Fig.1 (b))**.The automated approach enabled consistent, reproducible 3D region-of-interest (ROI) delineation across all patient scans, which is fundamental for reliable radiomic feature extraction. By eliminating inter-observer variability associated with manual segmentation, it ensured that radiomic features—including first-order statistics, texture patterns, and shape descriptors—were derived from standardized volumetric masks including GLCM, GLRLM, GLSZM, GLDM, and NGTDM. This consistency is critical for radiomic analysis, as feature values are highly sensitive to segmentation boundaries. Furthermore, automation allowed for efficient batch processing of the entire cohort, facilitating the extraction of thousands of quantitative imaging features that would be impractical to obtain manually.

### Extraction of anatomy features

Building on previous studies highlighting the relevance of local anatomy for endoleak risk assessment^[32–34]^, we developed a semi-automatic system to extract key morphological parameters from the aneurysm lumen model. In this study, we developed a system for semi-automatic extraction of morphological parameters to efficiently obtain local anatomical features after deriving the aneurysm lumen as a geometric model **(Fig.1 (c) i)**^[35]^. Our semi-automatic approach was implemented as follows: Following automated segmentation, the lumen surface mesh was loaded into 3D Slicer with the Segment Geometry module. An initial centerline was automatically computed using the Voronoi diagram method. An interventional radiologist then visually verified the centerline path and made manual adjustments at branching points if needed (required in approximately 8% of cases). After centerline validation, cross-sectional contours were automatically generated perpendicular to the centerline at 1-mm intervals. Diameter, area, and other morphological parameters were then automatically computed from these contours, with results displayed in an interactive table for final verification by the clinician.The measured features included:

- 1D parameters: the aortic plane of the lower margin of the renal artery(*P*_1_, *D*_1_), the abdominal aorta of the distal normal segment of the proximal aneurysm neck(*P*_2_, *D*_2_), the bifurcation plane of the aorta(*P*_3_, *D*_3_), the right(*P*_4_, *D*_4_) and left iliac artery starting plane(*P*_5_, *D*_5_) the maximum diameter of the AAA(*D*_*max*_). The *Proximal neck length* of aneurysm, the circumference of the plane where the maximum diameter of AAA located (*C*_*max*_), the center-line curvilinear length of AAA (*L*), the center-line straight length of AAA (*L*’) and the aneurysm height (*l*);
- 2D parameters: *Asymmetry*, *Tortuosity*, *Saccularization index*, *Width* − *height ratio*, *Deformation ratio* and *Infrarenal angulation* (*β*);
- 3D parameters: AAA volume and surface area.

We measured a total of 11 1D features, 6 2D features, and 2 3D features. Detailed measurement procedures are provided in the **Supplementary Material (Fig.S1)**.

Global morphological features are important for predicting vascular disease progression and postoperative outcomes^[36–40]^. The Statistical Shape Model (SSM) is a key method for analyzing overall morphological variations. Previous studies have shown that SSM can effectively classify arterial conditions and predict aortic aneurysm growth when combined with regression methods. In this study, the core workflow of global morphological features extraction comprises 2 sections: registration **(Fig.1 (c) ii)** and feature extraction **(Fig.1 (c) iii)**^[41]^. Detailed procedures are provided in the **Supplementary Material (Fig.S2)**.

### Extraction of hemodynamic features

Endoleaks after EVAR may result from abnormal hemodynamics, potentially leading to stent migration or seal failure. Preoperative hemodynamic parameters—including TAWSS, OSI, and RRT—are predictive of endoleak risk, as low WSS promotes flow stagnation and thrombosis, high OSI exacerbates endothelial dysfunction, and prolonged RRT increases coagulation^[42,43]^.

CFD simulations based on patient-specific 3D models enable quantitative hemodynamic analysis by solving Navier-Stokes equations under physiological boundary conditions. Key steps include vascular reconstruction from CTA, application of clinical boundary conditions, and numerical solution with refined meshing for accuracy. Despite its value in correlating hemodynamics with AAA progression, CFD remains limited in clinical use due to high computational cost, complex preprocessing, and long modeling time^[44,45]^.

CFD simulations discretize fluid domains into millions of nodes and tens of millions of elements for iterative equation solving. Despite hardware/algorithmic advances, single AAA cases still require tens of hours to days. This study develops an semi-automated CFD framework for rapid AAA simulation **(Fig.1 (d))**, which included the integratedpipeline model preprocessing; intelligent boundary condition assignment; high-performance numerical solving; Batch hemodynamic parameter extraction. Finally, established a standardized hemodynamic database. Detailed procedures are provided in the **Supplementary Material (Fig.S3).**

### Six types of ML models

Six ML models were subsequently developed: SVM, XGBoost, and Logistic Regression algorithms each implemented in two configurations – a base branch fusing history and morphological features (H+M), and an enhanced branch incorporating history, morphological, and Radiomic features (H+M+R) – to decode the mapping between 1D input features and endoleak outcomes **(Fig.1 (e) Strategy 1)**. SHAP (SHapley Additive exPlanations) analysis was then applied to quantify feature contributions and evaluate feature-endoleak associations.

To improve model performance, computational efficiency, and interpretability, feature selection was performed using Lasso regression. Features with an absolute correlation below 0.05 with the target were removed to eliminate noise and weak predictors. Additionally, to reduce multicollinearity, features with correlation coefficients above 0.8 were excluded, retaining only relevant and non-redundant predictors.

### Four DL methods based on Point Cloud Neural Networks

Using Python’s trimesh library, we extracted vertex coordinates (X-Y-Z) from STL-format AAA lumen models and stored them as NumPy Zip Archive (NPZ) point cloud datasets. The data was normalized to [0,1] range, resulting in 381 standardized AAA lumen point clouds.

Hemodynamic parameters (TAWSS, OSI, RRT) were computed for each patient using a semi-automated CFD pipeline and mapped onto corresponding mesh nodes. This enriched the original spatial coordinates (X, Y, Z) with three biomechanical features, converting each point into a 6D vector (X, Y, Z, TAWSS, OSI, RRT). The resulting dataset comprised 381 biomechanically augmented AAA point clouds stored in .npz format.

Finally, clinical history data and Radiomic features were integrated into the neural network architecture. The overall workflow is illustrated in **Fig.3.**

**Figure 3.**
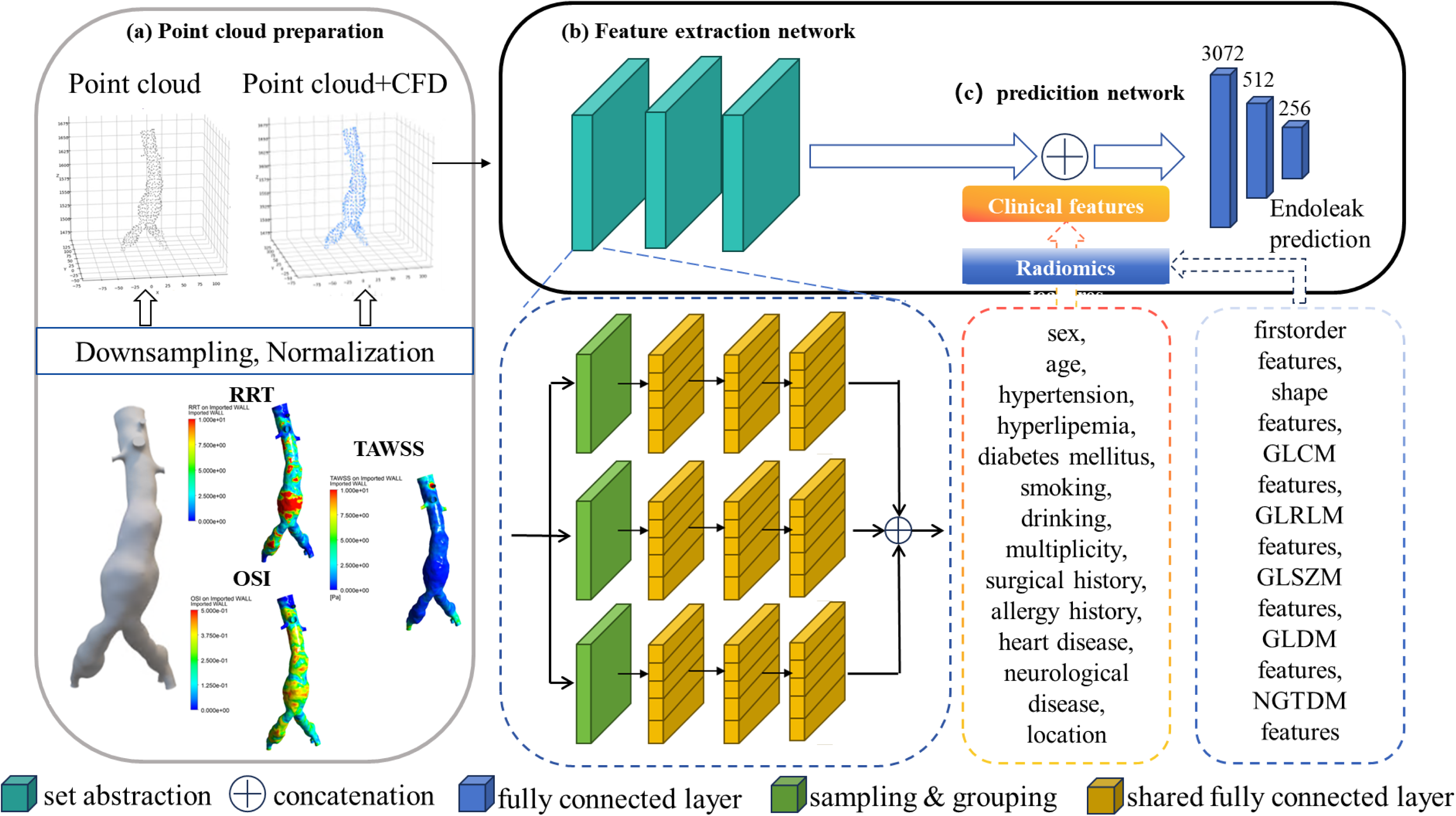
(a) Point cloud data preparation.The post-CFD point clouds incorporated three biomechanical features, transforming each data point from a 3-dimensional vector (X, Y, Z) into a 6-dimensional vector (X, Y, Z, TAWSS, OSI, RRT). (b) Feature extraction network structure. The feature extraction network comprises three set abstraction levels. The first two levels each consist of three parallel branches with identical sampling rates but distinct grouping radii. (c) Endoleak prediction network structure. An initial FC layer maps the 3072D features to 512D (followed by BN, ReLU, and 0.4 dropout), a subsequent FC layer reduces it to 256D (followed by BN, ReLU, and 0.5 dropout), and a final FC layer projects onto the class dimension space for probabilistic prediction via a sigmoid activation.

The point cloud feature extraction network employs three set abstraction levels. The first two levels each contain three parallel branches with identical sampling rates but different grouping radii. Each branch uses sampling and grouping layers to divide the point cloud into local regions, followed by a three-layer fully connected network with cross-channel weight sharing to extract regional features. Multi-scale morphological characteristics are captured by concatenating features from all branches. The final level uses a single branch to integrate all features into a global 1024-dimensional descriptor. Through iterative sampling, grouping, and feature extraction, the network hierarchically captures features from local to global scales.

After incorporating hemodynamic parameters, each network processes a 4D point cloud (XYZ + one hemodynamic feature). Three independent feature extraction networks—each with three set abstraction levels—process the data. The first two levels use three parallel branches (varying radii, same sampling rate) to extract multi-scale features via sampling, grouping, and shared weight fully connected (FC) layers. The final level produces a 1024-dimensional global descriptor per network. The three resulting feature vectors are concatenated into a 3072-dimensional vector, which is used as input to the endoleak prediction network.

The prediction head for this fused input utilizes a similar FC stack: an initial FC layer maps the 3072D features to 512D (followed by Batch Normalization (BN), ReLU, and 0.4 dropout), a subsequent FC layer reduces to 256D (followed by BN, ReLU, and 0.5 dropout), and a final FC layer projects onto the class dimension space for probabilistic prediction via a sigmoid activation. Subsequently, we constructed four distinct DL architectures: (a) A PCNN integrating only clinical history information; (b) A RA-PCNN model embedding Radiomic features within the PointNet architecture; (c) A CFD-PCNN model integrating hemodynamic parameters; (d) An integrated RA-CFD-PCNN model achieving cross-modal feature coupling, as illustrated in **Fig.1 (e) Strategy 2.** This progressive feature fusion strategy systematically explores the synergistic predictive mechanisms of incorporating features from different modalities on endoleak occurrence.

### Statistical analysis, performance metrics, and reporting

For the two types of prediction models, the performance evaluation metrics mainly include Accuracy (Equation 3), Precision(Equation 4), Recall (Equation 5), and F1 Score(Equation6) are calculated as follows:

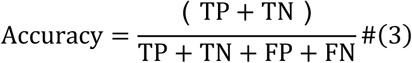

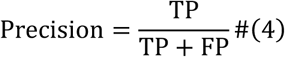

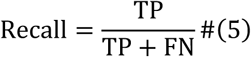

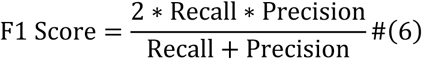

The 381 AAA cases were split into a training-validation set (n=286) and a test set (n=95). Five-fold cross-validation was applied during training. Ten models-six traditional ML and four DL methods-were evaluated on the test set using accuracy, precision, recall, F1-score, and AUC.

This study compared the predictive performance of ML and DL models using multiple evaluation metrics across two distinct dimensions.

Horizontal Dimension: This study compared traditional ML models (SVM, XGBoost, Logistic Regression) with a semi-end-to-end PCNN-based DL architecture. The analysis focused on differences in feature extraction, decision boundary formation, and endoleak prediction performance. Emphasis was placed on evaluating how the inclusion of Radiomic features—beyond morphological and clinical data — affects predictive accuracy, contrasting inherently interpretable models with black-box approaches.

Vertical Dimension: This dimension systematically assesses the influence of progressively enriching feature combinations on model performance. Performance changes were compared across distinct feature integration stages: **Stage 2.1:** History + Morphology (H+M, PCNN group); **Stage 2.2:** Addition of Radiomic (H+M+R, RA-PCNN group); **Stage 2.3:** Intergration of Hemodynamics (H+M+CFD, CFD-PCNN group); and **Stage 2.4:** Inclusion of All Parameters (H+M+R+CFD, RA-CFD-PCNN group). This longitudinal analysis progressively quantifies the performance evolution of the DL prediction models across these four distinct feature integration phases.

The study performed an exploratory analysis using the best-performing ML and DL models to predict endoleak risk in all AAA patients. Based on predictions, patients were classified into endoleak and non-endoleak groups. Kaplan-Meier survival curves were plotted, and group differences were assessed with the log-rank test. Hazard ratios (HR) were calculated using a Cox proportional hazards model after verifying proportional hazards assumptions.

## Results

### Feature extraction and selection

Clinical historical characteristics including gender, age, smoking status, alcohol consumption, hypertension, hyperlipidemia, diabetes mellitus, neurological disorders, cardiovascular diseases, urological diseases, respiratory diseases, surgical history, and allergy history were systematically recorded during treatment, with detailed statistical information presented in **Table 1**.

**Table 1.**
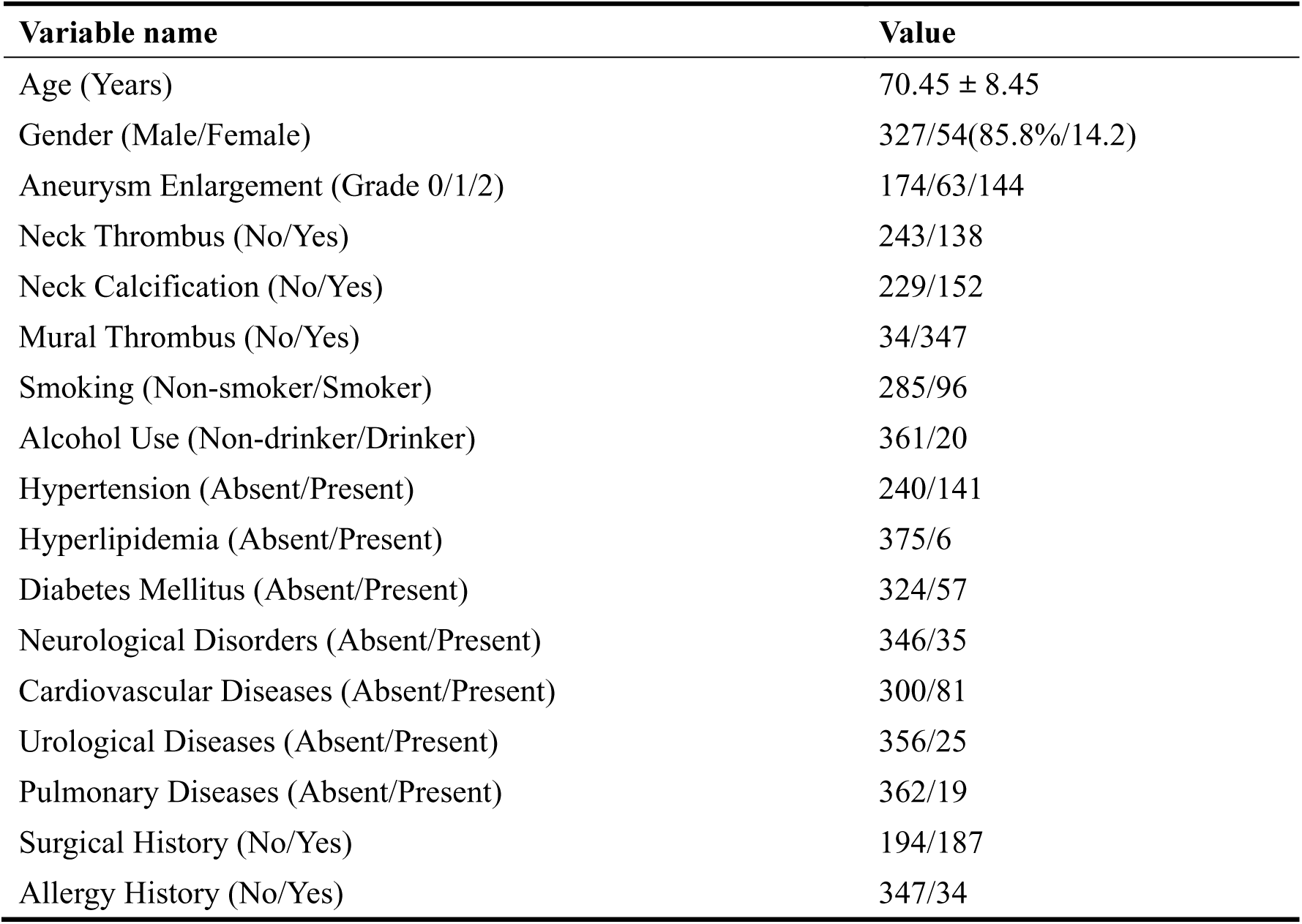
Clinical historical features of AAA patient.

This study developed a semi-automated AAA local anatomical feature measurement system, extracting 11 one-dimensional (1D), 6 two-dimensional (2D), and 2 three-dimensional (3D) parameters. Results are summarized in **Table 2**. The global morphological parameters are denoted as PC_i_(i=1,2,3…n), where n represents the total sample size. Each principal component PC_i_ satisfies: Mean (PC_i_)=0, std (PC_i_)=1.

**Table 2.**
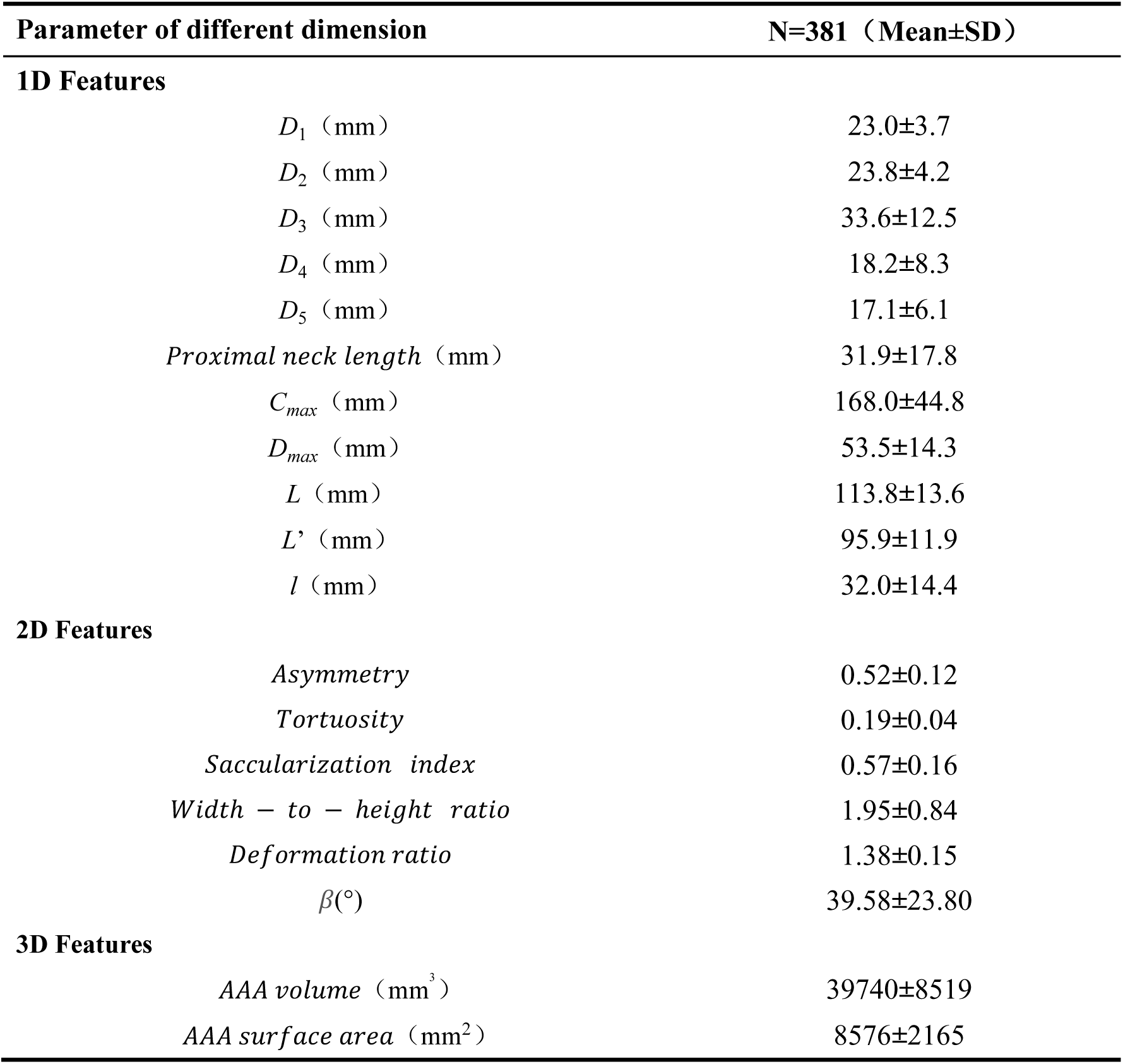
Statistical information of Local Anatomical Features.

| Parameter of different dimension | N=381 (Mean±SD) |
| --- | --- |
| **1D Features** |  |
| $D_1$ (mm) | 23.0±3.7 |
| $D_2$ (mm) | 23.8±4.2 |
| $D_3$ (mm) | 33.6±12.5 |
| $D_4$ (mm) | 18.2±8.3 |
| $D_5$ (mm) | 17.1±6.1 |
| *Proximal neck length* (mm) | 31.9±17.8 |
| $C_{max}$ (mm) | 168.0±44.8 |
| $D_{max}$ (mm) | 53.5±14.3 |
| $L$ (mm) | 113.8±13.6 |
| $L'$ (mm) | 95.9±11.9 |
| $l$ (mm) | 32.0±14.4 |
| **2D Features** |  |
| *Asymmetry* | 0.52±0.12 |
| *Tortuosity* | 0.19±0.04 |
| *Saccularization index* | 0.57±0.16 |
| *Width – to – height ratio* | 1.95±0.84 |
| *Deformation ratio* | 1.38±0.15 |
| $\beta(^{\circ})$ | 39.58±23.80 |
| **3D Features** |  |
| *AAA volume* (mm<sup>3</sup>) | 39740±8519 |
| *AAA surface area* (mm<sup>2</sup>) | 8576±2165 |

Hemodynamic features—including TAWSS, OSI and RRT—were calculated as described in **Supplementary Material (Fig.S3)**. Spatial means of these parameters were subsequently derived, TAWSS at 1.97±0.39 Pa, OSI at 0.31±0.04, and RRT at 6.39±0.68 Pa⁻¹.

In the Methods section, feature selection was performed using Lasso regression. The selected anatomical parameters included *D*_3_, *D*_4_, *D*_5_, *L*’, *Asymmetry*, *Tortuosity*, *Width height ratio*, *Deformation ratio*, and *β*; concurrently, 4 relevant Radiomic features were identified: Least Axis Length, Maximum 3D Diameter, Minor Axis Length, and Surface Area, yielding a total of 9 morphological parameters and 4 Radiomic features for subsequent analysis.

### Endoleak Prediction Using ML Models

In this study, the dataset was partitioned with a training-validation set to test set ratio of 3:1. During model training, five-fold cross-validation was employed on the training-validation set, and the model achieving the highest AUC value was selected for final evaluation on the test set. The training-validation set consists of 76 endoleak cases and 210 non-endoleak cases, while the test set contains 30 endoleak cases and 65 non-endoleak cases. This partitioning demonstrates an approximately 2.76:1 ratio of non-endoleak to endoleak samples in both the training-validation and test sets, maintaining consistent class distribution across splits for model development and evaluation. The total dataset comprises 106 endoleak cases and 275 non-endoleak cases.

When utilizing only local anatomy, global morphology, and clinical history as input features, the performance of XGBoost, Logistic Regression, and SVM during training, assessed via 5-fold cross-validation, is detailed in Table 3. The prediction results of the test set is detailed in Table 4. XGBoost (AUC = 0.711, accuracy = 0.729) and Logistic Regression (AUC = 0.706, accuracy = 0.635) both outperformed the SVM (AUC = 0.553, accuracy = 0.656) in predictive performance on the test set.

**Table 3.** The 5-fold cross-validation results of Type I (morphological + history) machine learning approach.

|  | XGBoost | Logistic regression | SVM |
| --- | --- | --- | --- |
| Fold 1 | 0.734 | 0.700 | 0.524 |
| Fold 2 | 0.744 | 0.607 | 0.608 |
| Fold 3 | 0.723 | 0.771 | 0.456 |
| Fold 4 | 0.671 | 0.642 | 0.582 |
| Fold 5 | 0.706 | 0.684 | 0.309 |
| Mean | 0.716 | 0.680 | 0.496 |

**Table 4.**
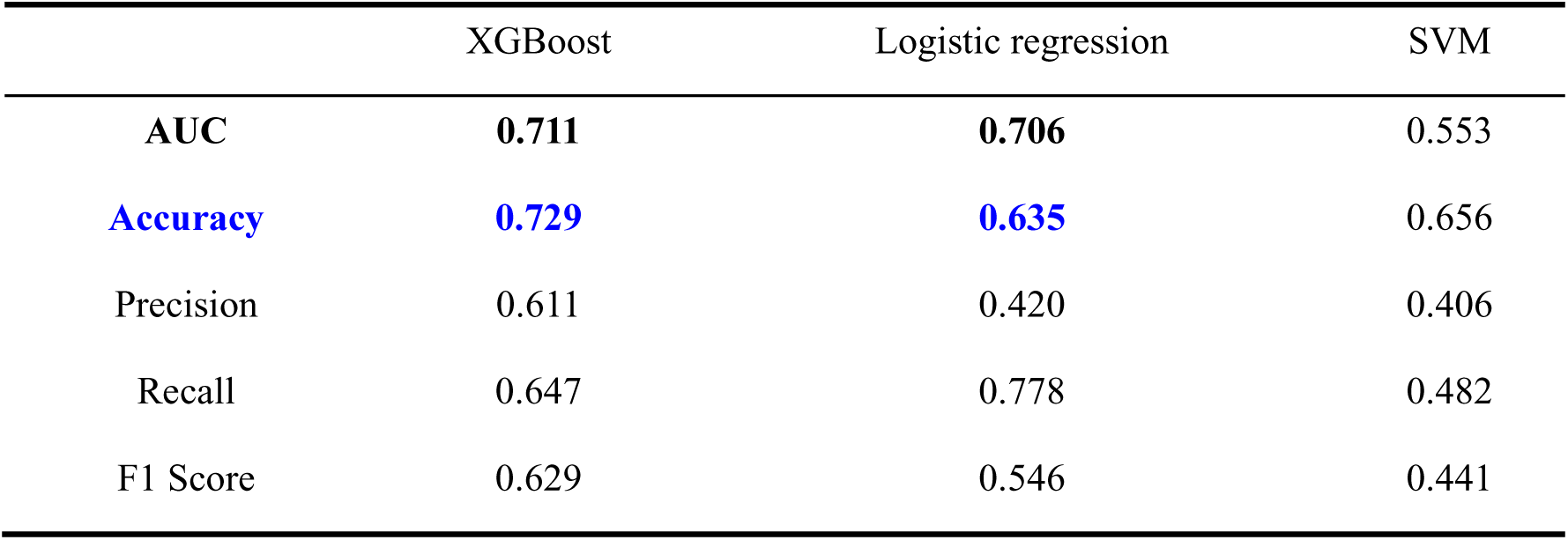
The prediction results of Type I (morphological + history) machine learning approach on the test set.

Following the incorporation of Radiomic features, the 5-fold cross-validation results are presented in Table 5. On the test set, the predictive outcomes of Type II ML approaches are summarized in Table 6. Notably, compared to models without Radiomic features, the XGBoost and Logistic Regression approaches demonstrated relative AUC improvements of 8.72% and 4.00%, respectively. Nevertheless, both AUC values remained below the 0.8 threshold.

**Table 5.** The 5-fold cross-validation results of Type II (morphological + history + radiomics) machine learning approach.

|  | XGBoost | Logistic regression | SVM |
| --- | --- | --- | --- |
| Fold 1 | 0.738 | 0.733 | 0.524 |
| Fold 2 | 0.738 | 0.684 | 0.608 |
| Fold 3 | 0.641 | 0.694 | 0.456 |
| Fold 4 | 0.660 | 0.665 | 0.418 |
| Fold 5 | 0.698 | 0.684 | 0.691 |
| Mean | 0.695 | 0.691 | 0.540 |

**Table 6.** The prediction results of Type II (morphological + history + radiomics) machine learning approach on the test set.

|  | XGBoost | Logistic regression | SVM |
| --- | --- | --- | --- |
| **AUC** | **0.773** | **0.734** | 0.605 |
| **Accuracy** | **0.719** | **0.729** | 0.552 |
| Precision | 0.533 | 0.541 | 0.340 |
| Recall | 0.800 | 0.690 | 0.630 |
| F1 Score | 0.640 | 0.606 | 0.442 |

The endoleak cohort comprised 28 patients with type I endoleak, 91 patients with type II endoleak, and an overlap cohort of 13 patients presenting with both sub-types; additionally, 275 patients were categorized in the non-endoleak group.

We employed the top-performing ML model on the test set, XGBoost, to predict endoleak classification. The prediction results are illustrated in **Fig. 4(a)-(b)**. With AUCs of 0.609 for type I endoleak and 0.708 for type II endoleak.

**Figure 4.**
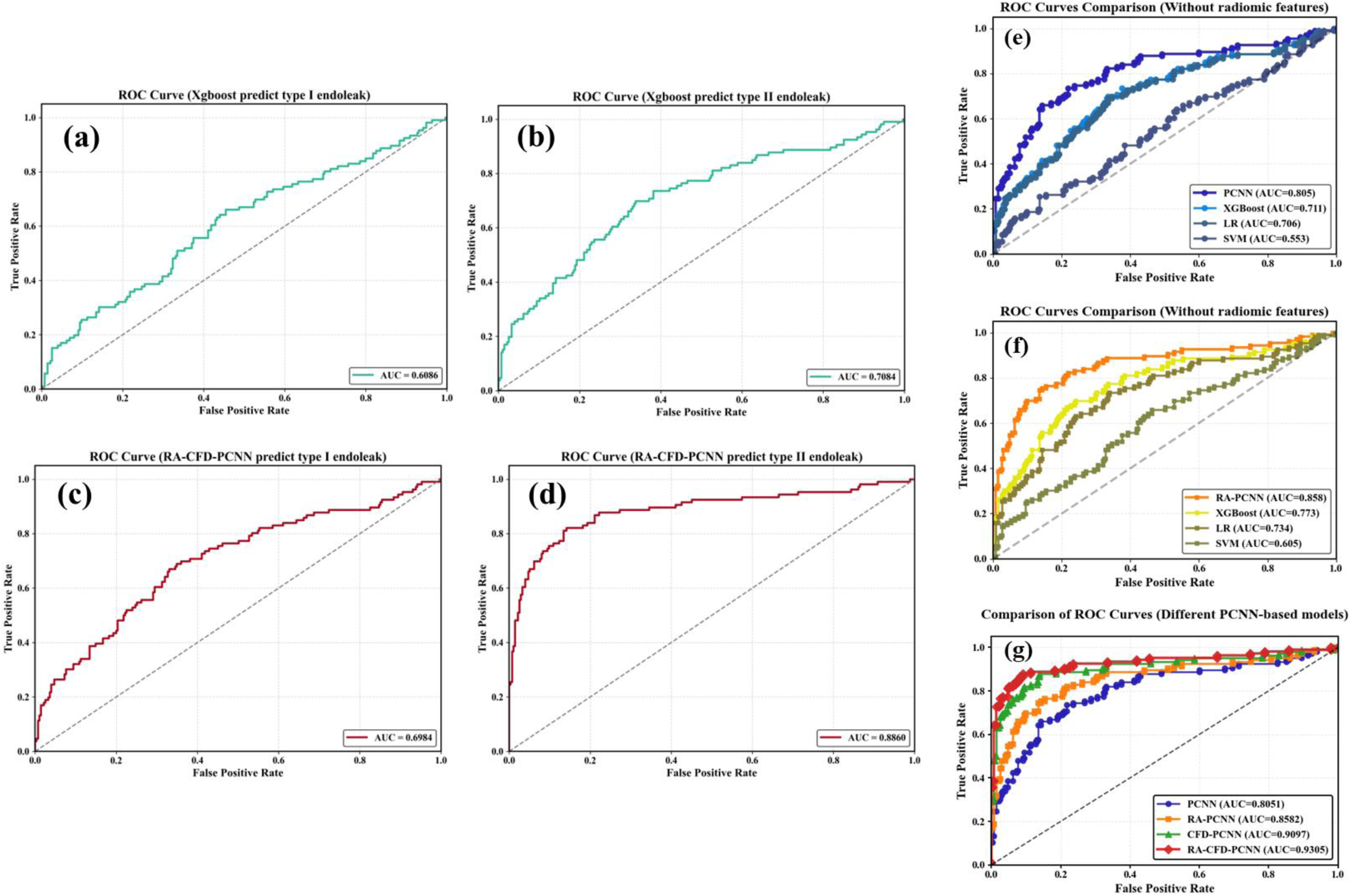
The predictive performance using ML and DL models. (a): The performance of type I endoleak prediction using XGBoost. (b): Type II endoleak prediction using XGBoost. (c): Type I endoleak prediction using RA-CFD-PCNN. (d): Type II endoleak prediction using RA-CFD-PCNN. (e): Cross-sectional comparison between machine learning models and deep learning models without radiomic features. (f): Cross-sectional comparison between machine learning models and deep learning models with radiomic features. (g): Differences in endoleak prediction performance of the 4 PCNN-based frameworks.

Using an XGBoost model for the prediction and classification of endoleaks, we conducted an analysis of feature importance and visualized the top 10 most significant features in **Fig. 5(a)-(c)**. In addition, we identified that PC_2_(the value of the second principal mode derived from SSM) consistently emerged as a statistically significant predictor across all three endoleak prediction tasks. This finding supports the rationale that extracting such global features via SSM from diverse AAA models substantially influences aneurysm development and can serve as an effective input variable for endoleak prediction. As the shape feature coefficient increases, the aneurysm exhibits a tendency toward unilateral expansion, manifesting primarily as an enlargement of the inferior sac. This progressive dilatation becomes more pronounced with higher feature values, as illustrated in **Fig. 5(d)**.

**Figure 5.**
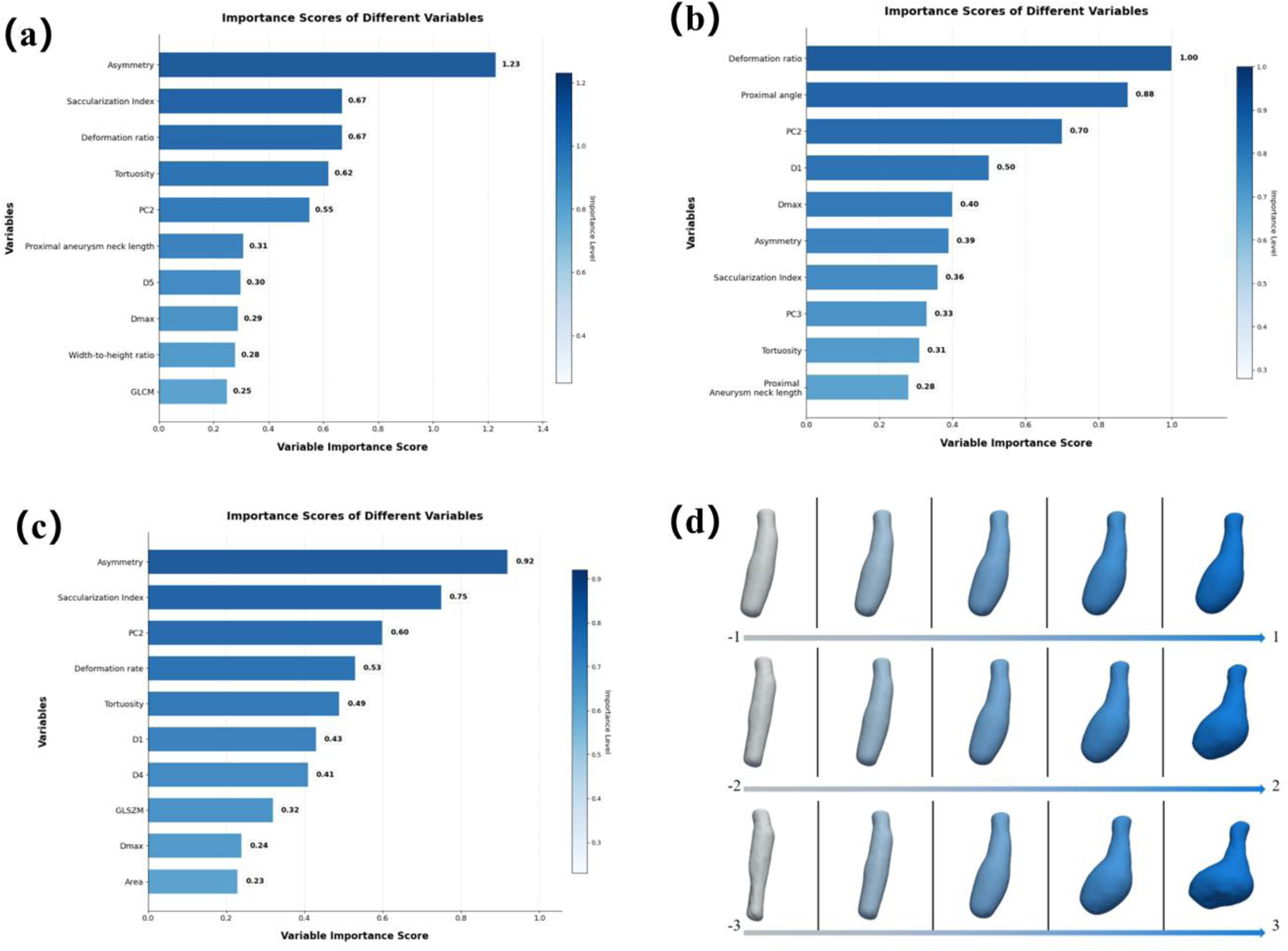
(a): The top 10 most important features during endoleak prediction using XGBoost. (b): The top 10 most important features during type I endoleak prediction using XGBoost. (c): The top 10 most important features during type II endoleak prediction using XGBoost. (d): Effects of alterations in PC2 on AAA morphology.

### Endoleak Prediction Using Point Cloud Neural Networks

The 5-fold cross-validation results during training for 4 PCNN-based prediction models are detailed in **Table 7**. Test set performance of the 4 PCNN frameworks is summarized in **Table 8**.

**Table 7.**
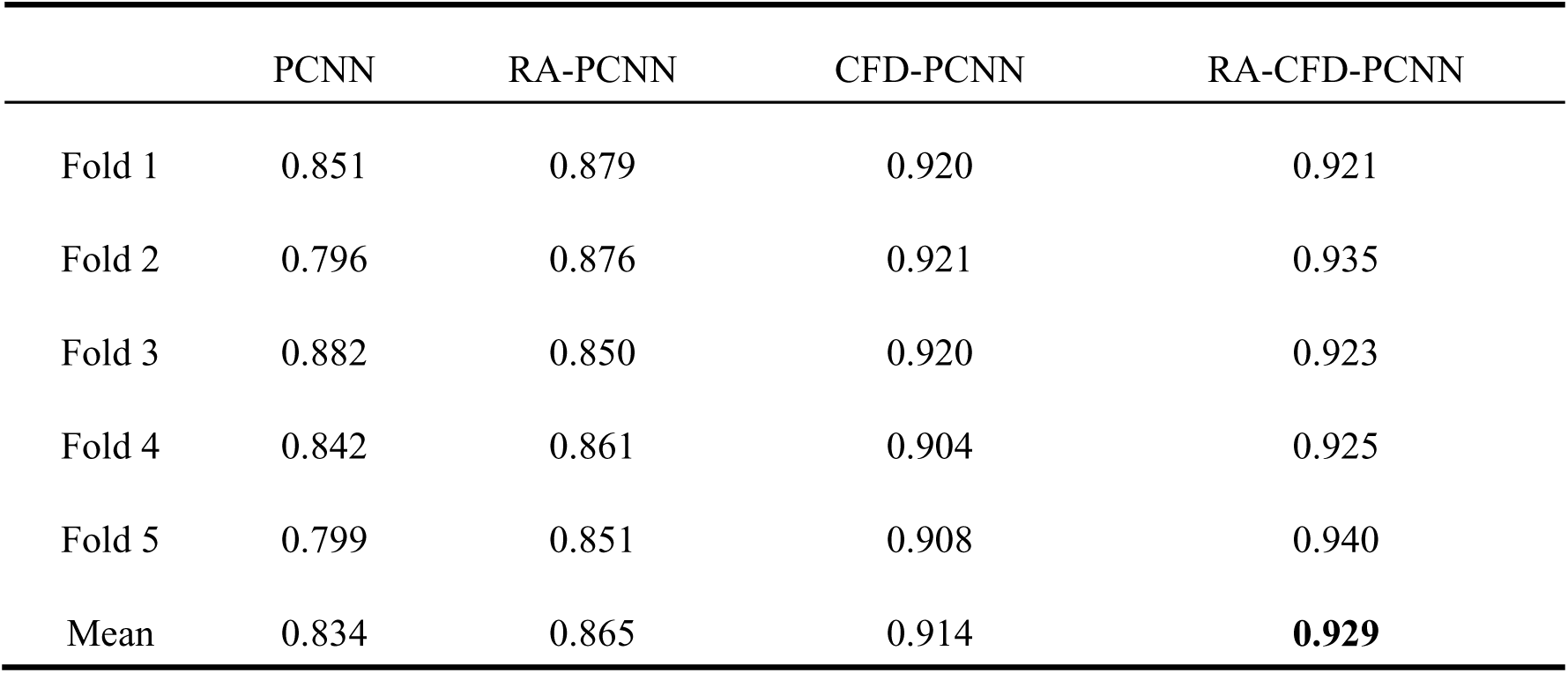
The 5-fold cross-validation results of 4 PCNN-based prediction models.

|  | PCNN | RA-PCNN | CFD-PCNN | RA-CFD-PCNN |
| --- | --- | --- | --- | --- |
| Fold 1 | 0.851 | 0.879 | 0.920 | 0.921 |
| Fold 2 | 0.796 | 0.876 | 0.921 | 0.935 |
| Fold 3 | 0.882 | 0.850 | 0.920 | 0.923 |
| Fold 4 | 0.842 | 0.861 | 0.904 | 0.925 |
| Fold 5 | 0.799 | 0.851 | 0.908 | 0.940 |
| Mean | 0.834 | 0.865 | 0.914 | **0.929** |

**Table 8.** The prediction results of 4 PCNN-based prediction models on the test set.

|  | PCNN | RA-PCNN | CFD-PCNN | RA-CFD-PCNN |
| --- | --- | --- | --- | --- |
| **\*AUC\*** | **0.805** | **0.858** | **0.910** | **0.930** |
| Accuracy | 0.803 | 0.829 | 0.866 | 0.900 |
| Precision | 0.642 | 0.672 | 0.710 | 0.788 |
| Recall | 0.660 | 0.755 | 0.877 | 0.877 |
| F1 Score | 0.651 | 0.711 | 0.785 | 0.830 |

\*Notes: RA = Radiomic features; CFD = Computational fluid dynamics features. The random guess baseline AUC (0.5016) reflects a 50.2%/49.8% class distribution. \*

Incorporating different feature combinations into the PCNN architecture differentially impacted predictive outcomes. While the baseline PCNN model achieved an AUC of 0.805, augmentation with Radiomic and CFD features elevated performance to 0.858 and 0.910, respectively. Notably, integrating all feature combinations increased the AUC to 0.930, demonstrating the efficacy of our selected feature sets for endoleak prediction.

Corresponding results for the top-performing DL model, RA-CFD-PCNN model, demonstrated AUCs of 0.700 for type I and 0.886 for type II endoleaks **(Fig. 4(c)-(d))**. Although RA-CFD-PCNN outperformed XGBoost (AUCs of 0.609 for type I endoleak and 0.708 for type II endoleak) **(Fig. 4(a)-(b))**, the predictive performance for type I endoleak remained suboptimal in both models, with neither AUC exceeding 0.7. This limitation may be attributed to the relatively late onset of type I endoleaks compared to type II, coupled with the small cohort size (only 28 positive cases), resulting in class imbalance and consequently diminished predictive efficacy.

The survival analysis similarly showed no significant difference in time-to-event rates between groups classified by either model as “Type I Endoleak” or “No Type I Endoleak” (P > 0.05) **(Fig. 6(c)-(d))**. However, the difference in survival rates between the “Endoleak” and “Non-Endoleak” groups classified by the PCNN model was more significant (HR 4.84, 95% CI: 2.74–8.53; P < 0.0001) than that classified by the XGBoost model (HR 3.21, 95% CI: 1.69–6.13; P < 0.0001) **(Fig. 6(a)-(b))**. In the prediction of type II endoleaks, the PCNN model also performed better (HR 5.24, 95% CI: 2.83–9.72; P < 0.0001) than the XGBoost model (HR 2.43, 95% CI: 1.28–4.60; P < 0.0001) **(Fig. 6(e)-(f)).**

**Figure 6.**
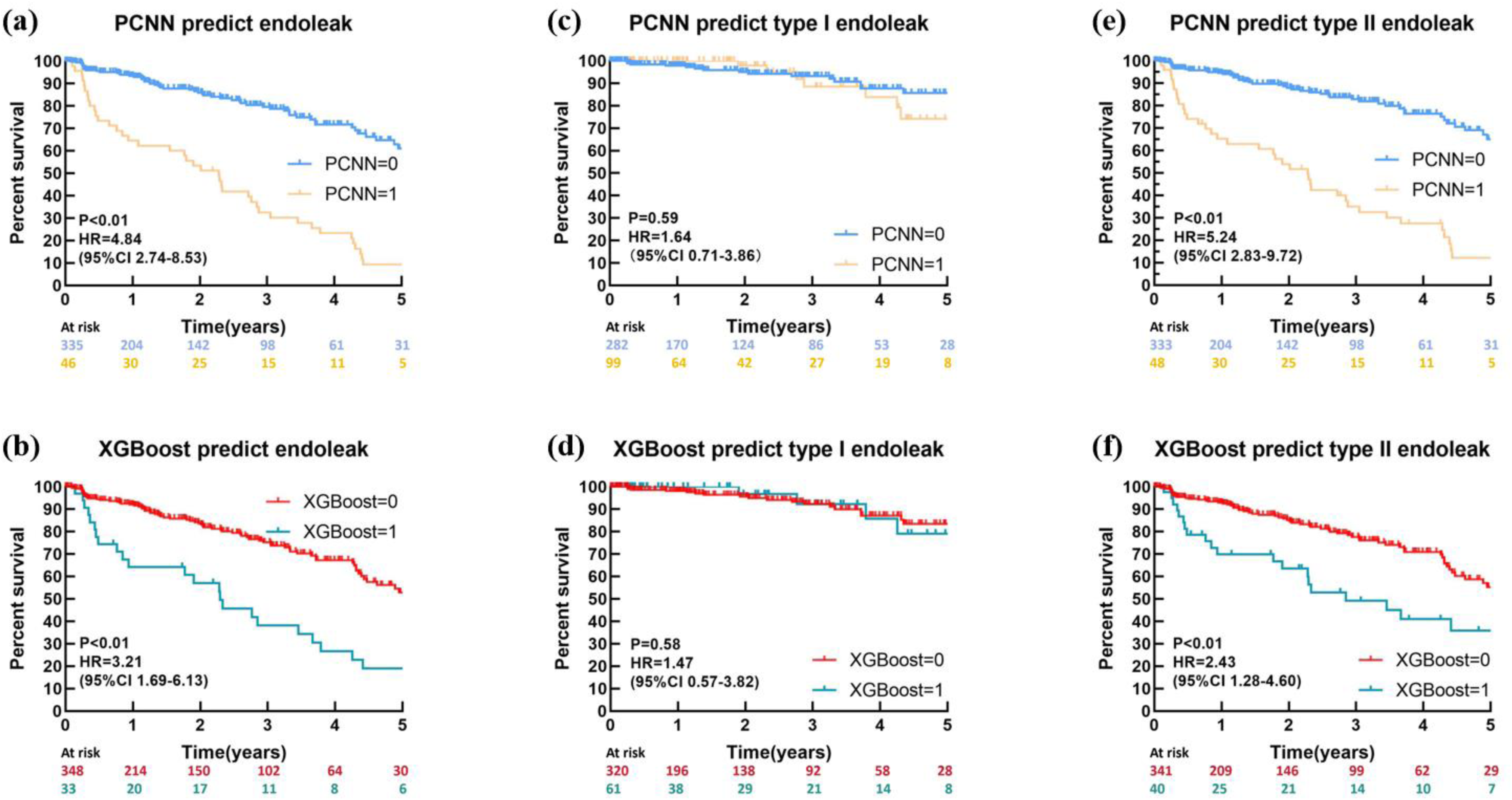
Kaplan-Meier curves based on machine learning and deep learning models for patient classification in different endoleak prediction tasks.Hazard ratios (HR) were calculated by fitting a Cox proportional hazards model. P-values are calculated using the log-rank test. (a):KM curves for the “Endoleak” and “Non-Endoleak” groups classified based on PCNN prediction results.“PCNN=0” indicates the absence of endoleak.(b):KM curves for the “Endoleak” and “Non-Endoleak” groups classified by XGBoost.“XGBoost=0” indicates the absence of endoleak. (c) :KM curves for the “Type I Endoleak” and “No Type I Endoleak” groups classified by PCNN. (d) :KM curves for the “Type I Endoleak” and “No Type I Endoleak” groups classified by XGBoost. (e) : KM curves for the “Type II Endoleak” and “No Type II Endoleak” groups classified by PCNN. (f) :KM curves for the “Type II Endoleak” and “No Type II Endoleak” groups classified by XGBoost.

We compared all three ML methods and the PCNN prediction model without incorporating Radiomic features, with the results shown in **Fig. 4(e)-(f)**. Subsequently, we compared ML methods with the RA-PCNN prediction model after the integration of Radiomic features, as demonstrated in **Fig. 4(g)**. It can be observed that both with and without the inclusion of Radiomic features, the AUC values achieved by the PCNN and RA-PCNN prediction models were significantly higher than those of the ML models.

To investigate the reason for this performance discrepancy, we further examined the model architecture. During the point cloud data processing stage, normalization is a critical step for eliminating dimensional discrepancies and enhancing model convergence efficiency. A three-level feature abstraction layer (incorporating down-sampling and non-linear activation) progressively extracts a 1024-dimensional high-level feature vector. This process employs hierarchical convolutional kernels to aggregate local geometric features, ensuring the final output vector preserves the topological structure and key morphological information (e.g., surface curvature, edge gradients) of the original model, thereby providing a discriminative foundation for subsequent classification

We conducted Principal Component Analysis (PCA) on the feature vector space and visualized the first three principal components. As shown in **Fig. 7**, the eigenvalue distributions and corresponding 3D point clouds reveal distinct patterns between the two groups: the endoleak group tends to exhibit higher values along the first and third principal components but lower values along the second compared to the non-endoleak group. The probability density curves further illustrate this separation: endoleak samples (blue) display a unimodal, concentrated distribution within [0, 2], while non-endoleak samples (orange) show a broader, multimodal distribution across [−2, 2]. This highlights the strong discriminative ability of the first principal component (PCA1) in characterizing endoleaks. Moreover, the clear spatial separation between the two groups in the 3D visualization intuitively demonstrates the class separability enabled by these components.

**Figure 7.**
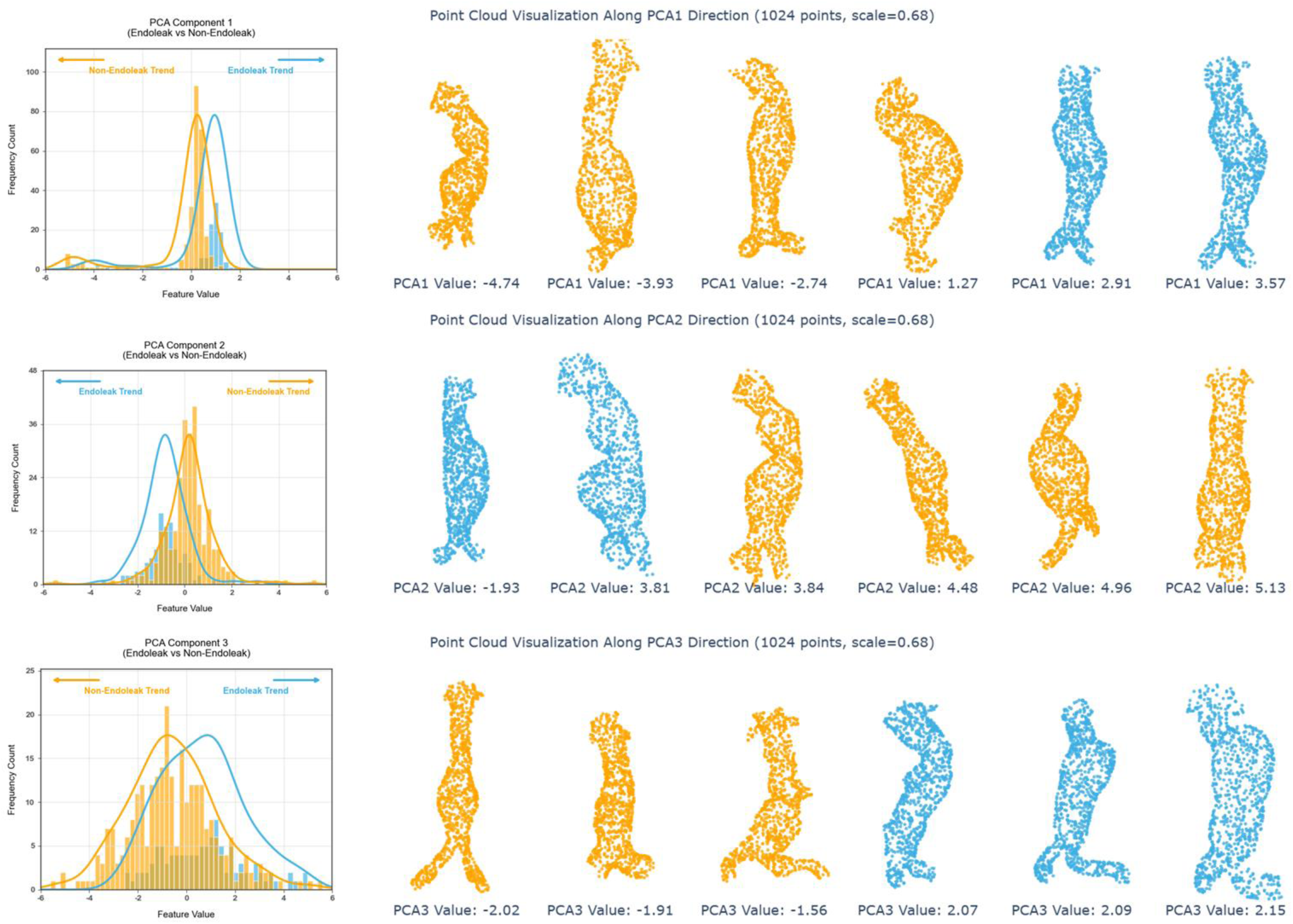
Three principal components analysis and 3D point cloud visualization. Along the 1st and 3rd principal components, the endoleak group-- blue label --tends to achieve higher values. Along the 2nd principal components, the non-endoleak group-- orange label -- tends to achieve higher values.The probability density curves further illustrate this separation: the curves corresponding to the endoleak group and the non-endoleak group exhibit distinct differences in both shape and position.

These PCA findings suggest that the PCNN model (AUC = 0.805), via its hierarchical feature learning, effectively captures morphological patterns in the latent space that align with these principal components. In contrast, traditional ML methods—with AUCs of 0.711, 0.706, and 0.553—remain limited by their dependence on handcrafted features, and are thus unable to adequately represent such complex, nonlinear geometric structures.

Subsequently, we compared the impact of incorporating different feature combinations on the predictive performance of the PCNN model. The results are shown in **Fig. 4(g)**. The inclusion of Radiomic features increased the AUC by 6.58%, while the addition of biomechanical features— specifically TAWSS, OSI, and RRT, which are significantly associated with WSS—led to a 13.04% improvement in AUC compared to the results of PCNN.

The most substantial performance improvement observed with the incorporation of CFD features can be explained through three primary mechanisms: (1) Psychophysiology relevance: Reduced TAWSS directly reflects blood flow stagnation, promoting thrombosis; elevated OSI indicates flow direction disorder predisposing to endothelial dysfunction; prolonged RRT is closely associated with platelet aggregation. These three parameters collectively form a “hemodynamic risk triad” demonstrating strong causal relationships with endoleak development; (2) Feature Stability: In 5-fold cross-validation, CFD-PCNN exhibited significantly lower standard deviation (0.007) compared to RA-PCNN (0.012) **(Table 7)**, indicating CFD features are less affected by data partitioning. This can be attributed to the inherent physical conservation laws governing hemodynamic parameters, which are derived from numerically solving the Navier–Stokes equations. As a result, these features offer a more physiologically grounded and mechanically consistent representation of blood flow behavior, making them more robust than purely data-driven Radiomic characteristics. (3) Clinical Interpretability: As shown in **Table 8**, CFD-PCNN’s advantages in recall (0.877 vs. RA-PCNN’s 0.755) and F1-score (0.785 vs. 0.711) are particularly prominent, demonstrating enhanced detection capability for positive cases. This originates from parameters like TAWSS directly identifying low shear stress regions, which are precisely the areas predisposed to aneurysm expansion and endoleak formation.

The integration of both Radiomic features and CFD features yielded optimal performance (AUC=0.930), demonstrating a clear synergistic effect of multimodal data fusion

This improvement suggests that the two feature types provide complementary and orthogonal information: Radiomic features primarily capture anatomical and structural properties, while CFD-derived parameters quantify functional hemodynamic status. By combining morphological and physiological perspectives, the model achieves a more comprehensive characterization of the pathophysiological mechanisms in endoleak development. Specifically, it reflects how the coupling of vascular geometric abnormalities with aberrant hemodynamic conditions collectively contributes to an increased risk of endoleaks.

## Discussion

Recent studies have begun to explore AI approaches for predicting the risk of postoperative complications following EVAR. For instance, Karthikesalingam et al. investigated the use of an artificial neural network (ANN) to predict complications and mortality after EVAR. Their retrospective analysis of 761 EVAR patients from two UK medical centers (2004–2010) demonstrated that the ANN model could effectively stratify patients into risk groups. The results showed significant differences in 5-year outcomes between high- and low-risk groups: aortic complications (95.9% vs. 67.9%), limb complications (99.3% vs. 92.0%), and mortality (87.9% vs. 79.3%) (p < 0.001)^[46]^. However, this approach combined various complications into a broad composite outcome and did not enable specific risk stratification for endoleaks. Lee et al. applied ML to identify strong correlations between Type I endoleaks and specific anatomical features of AAAs, particularly aneurysm neck length and diameter, as well as the suprarenal aortic angle. They also reported statistically significantly lower overall survival in patients with wide and very wide aortic necks, with 5-year survival rates of 86.1%, 84.8%, and 83.4% for normal, wide, and very wide necks, respectively^[47]^. Despite these insights, their methodology did not facilitate patient-specific prediction of endoleak risk. Wang et al. developed a DL model comparing multimodal frameworks based on morphological and Radiomic features to predict outcomes after EVAR. They found that a Radiomic model based on logistic regression showed superior predictive performance (AUC: 0.93, accuracy: 0.86, F1-score: 0.91)^[48]^. Nevertheless, this study did not incorporate the prediction of different endoleak subtypes. While these studies have progressively integrated clinical, morphological, and Radiomic features into predictive modeling, they have not yet incorporated biomechanical parameters. Furthermore, there remains a lack of a model capable of predicting different endoleak subtypes, representing a significant gap in the current research landscape. Therefore, in this paper, we investigated the feasibility of ML and DL methods for endoleak and its subtypes prediction using multimodal features and hemodynamic modeling. We evaluated the effectiveness of using hemodynamic features and examined whether it had a synergy effect for the prediction of endoleak or not. Our PCNN-based framework, which leverages the full 3D geometry through point-cloud representation and progressively fuses multimodal information and integrates CFD parameters by processing a 4D point cloud, where the three spatial coordinates (XYZ) are augmented with a hemodynamic feature. This is achieved through three independent feature extraction networks, each dedicated to processing this enhanced input. The PCNN-based architecture achieved substantially higher performance (AUC=0.81 for baseline PCNN; 0.86 with radiomics; 0.91 with hemodynamics; and 0.93 for the full RA-CFD-PCNN model) than ML methods. Notably, incorporating CFD-derived hemodynamic indices provided the largest incremental gain (+0.12 AUC over the baseline PCNN), highlighting the added clinical value of physiologically grounded features that are not captured by conventional anatomic descriptors alone. Beyond AUC, we also report values of accuracy, precision, recall, F1 score to compare the performance of different prediction models more comprehensively and further verify the necessity of incorporating CFD features.

In addition, point-cloud neural networks have recently been explored for vascular diseases prediction tasks beyond EVAR. Luo et al. combined a point cloud neural network (PCNN) with clinical variables and radiomics to predict cerebral aneurysm rupture, reporting an AUC of 0.83 in internal testing when incorporating parent-artery information on CTA dataset^[49]^. Zhou et al. similarly applied a PCNN framework with clinical features to predict distal aortic remodeling after thoracic endovascular aortic repair, achieving an AUC of 0.84^[50]^. These reported performances are comparable to our radiomics-enhanced RA-PCNN (AUC=0.86) and support the feasibility of point-cloud representations for capturing vascular morphology. Notably, in our ablation analysis, adding CFD-derived hemodynamic parameters yielded the largest incremental improvement (CFD-PCNN AUC=0.91), and the fully integrated RA-CFD-PCNN achieved the best discrimination (AUC=0.93), highlighting the added value of physiologically grounded hemodynamics for endoleak prediction after EVAR. This study establishes an integrated AI-driven framework for endoleak prediction after EVAR, leveraging computational simulations and multimodal data. Initially, automated segmentation extracted the aortic lumen and ILT, followed by Radiomic feature quantification. Subsequently, the segmentation masks were converted into geometric models, enabling algorithmic extraction of multidimensional anatomical and global morphological characteristics. A semi-automated CFD pipeline then derived high-dimensional hemodynamic parameters – including TAWSS, OSI, and RRT-which provide physiologically interpretable markers of flow stagnation and oscillation that are not captured by conventional diameter-based assessment alone. Compared with conventional patient-specific CFD workflows that are often difficult to translate into clinical routines due to extensive manual preprocessing (geometry cleaning, meshing, and case-by-case boundary condition setup) and operator dependence, our semi-automated implementation streamlines these steps by leveraging the automated 3D U-Net segmentation outputs and standardized scripts for mesh generation, boundary-condition assignment, and post-processing. This reduces manual operator time, improves reproducibility across patients, and enables cohort-scale hemodynamic characterization in a clinically feasible manner.

The semi-automated CFD approach offers significant clinical applicability by providing standardized hemodynamic assessment process that minimizes inter-operator variability, and enabling real-time visualization of hemodynamic parameters distribution that could assist in preoperative planning and device selection. The CFD outputs can be also summarized into a small set of scalar indices (e.g., spatially averaged TAWSS/OSI/RRT) together with intuitive wall maps, which may support clinically actionable use scenarios such as augmenting preoperative endoleak risk stratification and informing individualized follow-up necessity. We note that while the CFD solve itself remains computationally demanding, it is highly parallelizable and can be deployed as batch processing on hospital servers; ongoing work on surrogate modeling may further reduce turnaround time toward broader clinical adoption.

Results demonstrated that both PCNN and RA-PCNN outperformed conventional ML in cross-model comparisons. Notably, CFD-PCNN – enhanced with biomechanical features – achieved significant performance gains. The fully integrated RA-CFD-PCNN framework attained an AUC of 0.930. Performing patient-specific CFD simulations for all 381 cases is a computationally expensive task. To address this challenge, we innovatively developed a semi-automated CFD workflow for rapid hemodynamic feature extraction. First, we established an automatic segmentation system based on a 3D U-Net architecture to classify the lumen, ILT, and background. The segmented lumen geometry was used for CFD simulations, while the ILT region served for Radiomic feature extraction. This system significantly reduced the time required for AAA reconstruction and ROI delineation from raw CTA images. Next, to ensure a consistent computational framework, we applied boundary conditions derived from previously published in vivo measurements uniformly across all 381 AAA models^[51]^. Furthermore, given the relatively low pulsatility in the abdominal aortic region, cyclic vessel deformation was neglected^[16]^, which further streamlined the simulation process. Finally, custom scripts were developed to automatically extract key hemodynamic parameters—including TAWSS, OSI, and RRT—from simulation results generated in ANSYS CFX 2019 R3. Building upon this established framework, we are currently investigating the use of DL methods to optimize the CFD simulation component itself^[44,45,52–57]^. This refined workflow is well-suited for large-cohort hemodynamic analysis and helps overcome the limitations of small, high-cost, physics-based datasets for training deep neural networks.

We performed interpretability analysis on the constructed ML and DL models. Importantly, the SSM was built using preoperative CTA-derived AAA lumen geometries.

The rationale for introducing SSM is that conventional morphologic descriptors (e.g., maximum diameter, neck length/angulation, tortuosity indices) may not fully capture whole-aneurysm 3D shape complexity. In our workflow **(Supplementary Fig. S2),** AAA lumen surfaces were aligned via rigid and non-rigid registration to establish point-to-point correspondence across subjects, and PCA was then performed on the resulting deformation fields to obtain principal modes of global shape variation. Each patient receives a score along these modes (*PC*_*i*_), which can be used as compact global shape descriptors.

The results indicated that PC_2_ was a parameter with a significant impact on the outcome in the ML model, just as shown in **Fig. 5**. PC_2_ primarily quantifies a pattern of global asymmetry/unilateral sac expansion and increased tortuosity. Such abnormal morphology may complicate endograft delivery and positioning, increase the likelihood of suboptimal apposition at sealing zones, and alter flow patterns, thereby potentially predisposing to endoleak. While these global shape features reflect accumulated aneurysm remodeling along the AAA disease course, they remain directly available at the time of EVAR planning and may therefore be clinically actionable for preoperative risk stratification and procedural planning.

These insights could potentially aid clinicians in screening AAA patients at elevated risk for post-EVAR endoleaks based on global morphological characteristics. Indeed, several researchers have also applied SSM in biomedical studies of aortic aneurysm^[37,58,59]^. However, a universally reliable method for constructing SSMs across different anatomical regions remains lacking. Specifically, during the non-rigid registration process, landmark selection is both critical and challenging for establishing accurate point-to-point correspondence across models. This difficulty arises from anatomical variations between different organs^[60,61]^. In most cases, there is no consensus on the number of corresponding points required, and manual landmark alignment is prone to errors^[62]^. In this study, we adopted an automated approach to minimize repetitive manual intervention and proposed a robust framework that bypasses the need for organ-specific landmark identification during non-rigid registration. The results of the PCA performed on the DL model reveal that the superior performance of the PCNN over traditional ML methods stems from its ability to extract morphological features characterized by nonlinear geometric patterns in the latent space, which are strongly correlated with principal components. This suggests that as advanced methods for capturing complex morphological signatures—such as SSM and PCNN—continue to emerge, further substantial improvements can be expected in the field of endoleak prediction.

Also, we emphasize that SSM-derived *PC*_*i*_ should be interpreted as statistical shape descriptors and risk markers rather than direct causal drivers of endoleak; further multi-center validation is required to confirm their generalizability and to quantify incremental clinical utility beyond standard neck measurements.

In this study, the predictive model exhibited strong performance for Type II endoleaks but suboptimal accuracy for Type I. We suggest that this discrepancy stems from several factors. Foremost is the significant class imbalance, as the lower incidence of Type I endoleaks provided limited examples for the model to learn from. Large-scale clinical cohort studies have demonstrated that Type II endoleaks are the most frequent, with a reported incidence ranging from 14.0% to 25.3%, followed by Type I (0.6%-13.0%) and Type III (0.9%-2.1%) endoleaks. The occurrence of Type IV and Type V endoleaks is rare, each accounting for less than 1% of cases^[12]^. Additionally, the pathophysiological mechanisms underlying the two endoleak types also contribute to the performance disparity. Type II endoleaks, often stemming from retrograde flow through collateral vessels, may present more homogeneous and quantifiable morphological features. In contrast, Type I endoleaks result from complex and heterogeneous failure modes at the sealing zones (e.g., adverse neck anatomy, calcification), posing a greater challenge for pattern recognition. It is also possible that the current morphological and hemodynamic feature set, while effective for Type II, lacks the sensitivity to capture the subtle anatomical nuances critical for predicting Type I endoleaks. Future studies aimed at predicting all endoleak types should prioritize collecting larger multi-center cohorts to address class imbalance and explore more advanced features specifically designed to characterize proximal and distal seal zones.

Despite the successful development of our multimodal PCNN incorporating hemodynamic features, there are several limitations associated with the proposed method. **First**, the current study did not achieve a multi-class prediction task encompassing Type I, Type II, and combined Type I + II endoleaks simultaneously. Furthermore, beyond endoleaks, EVAR can lead to other complications such as stent graft migration and intimal injury^[63,64]^. Additionally, while this research focused specifically on postoperative AAA complication assessment, the proposed framework—pending improvements in efficiency and accuracy—holds potential for generalization to other vascular diseases. The interrelationships among different complications and across various vascular pathologies warrant further investigation. **Second**, the model was developed and validated using data from a single institution, and the absence of external validation from diverse clinical settings may limit its generalizability. Future studies should therefore aim to validate and extend our findings using larger sample sizes, prospective designs, multi-center longitudinal data, and more standardized data processing protocols. **Third**, this study did not incorporate other modal information, such as textual data. Future work should explore the integration of more diverse cross-modal and multi-dimensional data, including transient and long-term dynamic imaging, spatially distributed biomechanical patterns over the vascular wall, deep features from 2D images, Radiomic, and proteomics. To effectively harness such complex data, model architectures capable of enhanced feature extraction, correlation analysis across different feature types, and cross-modal feature alignment—such as Swin Transformer or Patch-GCN—should be adopted.

From a patient management perspective, the proposed framework is intended to function as a decision-support tool that complements, rather than replaces, existing clinical judgment. By integrating preoperative CTA-derived geometry with radiomic and hemodynamic descriptors, the model provides individualized risk estimates for post-EVAR endoleak that go beyond conventional diameter- or neck-based assessment alone. In clinical practice, such risk stratification may help identify patients who warrant closer postoperative surveillance, earlier follow-up imaging, or heightened peri-procedural attention, while potentially reducing unnecessary intensive imaging in patients predicted to be at low risk.

Importantly, the analytical validity of the framework is supported by its reliance on routinely acquired imaging data, standardized and largely automated feature extraction, and consistent performance across internal cross-validation and an independent test set. The strong contribution of CFD-derived hemodynamic parameters further suggests that incorporating physiologically meaningful flow information may enhance clinically relevant risk discrimination in EVAR patients. Nevertheless, we emphasize that prospective, multi-center validation is required before clinical deployment, and that the current model should be viewed as a tool to augment patient-specific risk assessment and inform individualized follow-up strategies rather than dictate management decisions.

The goal of medical AI is to augment the capabilities of surgeons, thereby enabling more precise and personalized diagnosis and treatment. By quantifying and learning from the expertise of highly skilled physicians, AI technology can provide decision support for junior doctors, thereby improving access to quality healthcare in underserved regions. From an individual patient’s perspective, AI-assisted tools empower surgeons to develop surgical plans tailored to specific anatomical and clinical characteristics, while also optimizing postoperative care protocols to significantly reduce the risk of complications. With the continued advancement of AI, clinicians will gain powerful intelligent support, fostering the sustainable development of healthcare systems.

## Conclusion

This study extracted clinical history, morphological, Radiomic, and hemodynamic parameters from 381 AAA patients and accomplished cross-modal feature fusion as well as endoleak prediction after EVAR using a PCNN-based approach. Through horizontal and vertical comparisons, the study demonstrated that the PCNN-based, multi-modal data-driven approach holds a significant advantage over traditional ML methods in predicting endoleaks. Furthermore, incorporating hemodynamic parameters into the PCNN framework substantially enhanced the model’s predictive performance, providing preliminary evidence for the necessity of integrating biomechanical features into predictive models. As the efficiency and accuracy of the predictive framework continue to improve, this methodology holds promise for generalization and application to the diagnosis of various other vascular diseases.

## Abbreviation

AAA: Abdominal Aortic Aneurysm
AI: Artificial intelligence
ANN: Artificial Neural Network
AUC: Area Under the Receiver Operating Characteristic Curve
BCA: Brachiocephalic Artery
BN: Batch Normalization
CA: Celiac Artery
CFD: Computational Fluid Dynamic
CNN: Convolutional Neural Network
CTA: Computed Tomography Angiography
D: Dimensional
DL: Deep Learning
DSA: Digital Subtraction Angiography
EVAR: Endovascular Aortic Aneurysm Repair
FC: Fully Connected
HR: Hazard Ratios
HU: Hounsfield Unit
ILT: Intraluminal Thrombus
LCCA: Left Common Carotid
LRA: Left Renal Artery
LSA: Left Subclavian
LEIA: Left External Iliac Artery
LIIA: Left Internal Iliac Artery
MACE: Major Adverse Cardiovascular Events
ML: Machine Learning
MRI: Magnetic Resonance Imaging
NPZ: NumPy Zip Archive
OSI: Oscillatory Shear Index
PCA: Principal Component Analysis
PCNN: Point Cloud Neural Network
RRT: Relative Residence Time
RA: Radiomic
ReLU: Rectified Linear Unit
REIA: Right External Iliac Artery
RIIA: Right Internal Iliac Artery
RRA: Right Renal Artery
ROC: Receiver Operating Characteristic Curve
SHAP: SHapley Additive exPlanations
SMA: Superior Mesenteric Artery
SSM: Statistical Shape Model
SVM: Support Vector Machine
TAWSS: Time-Averaged Wall Shear Stress
WSS: Wall Shear Stress
XGBoost: eXtreme Gradient Boosting

## Acknowledgments

This study was partially supported by the Youth Program of National Natural Science Foundation of China (No. 12402359), the National Natural Science Foundation of China (No. 82170493), the Fundamental Research Funds for the Central Universities (226-2025-00072), the Noncommunicable Chronic Diseases-National Science and Technology Major Project” (grant no. 2023ZD0504300).

## Data Availability Statement

The data will be shared at reasonable request to the corresponding author.

## Code Availability Statement

The code for this research is available via GitHub at https://github.com/fengyasi/PCNN

## Ethics declarations

The study was conducted in accordance with the Declaration of Helsinki, and approved by the Ethics Committee of Zhongshan Hospital, Fudan University (approval number, B2023-230). All including patients were informed about the nature of the study and gave their written informed consent. They all consented to the publishing of all clinical data, and other data included in the manuscript. The data underlying this article cannot be shared publicly due to ethical reasons.

---

**Figure S1** (a) Extraction of centerline. (b) Proximal aneurysm neck point 2 and distal aneurysm neck point 3. (c) Illustration of points and lines used for marking during extraction of local anatomical features. (d) Different locating planes. (e) Schematic diagram of 1D feature measurements. (f) Schematic Diagram of AAA volume and surface area quantification.

**Figure S2** (a) Pre-registration state. (b) Rigid registration in progress. (c) Rigid registration completed. (d) Non-rigid registration in progress. (e) Non-rigid registration completed. (f) Geodesic distance. (g) Intrinsic mean and tangent space.

**Figure S3** (a) Schematic diagram of AAA 3D anatomical model after processing. (b) Schematic diagram of meshing. (c) Magnitude and phase images within the cross-sectional segment. (d) 3D volumetric rendering depicting velocity profiles across 30 cardiac phases. (e) Descending thoracic aortic (SC) blood flow waveform over 30 temporal phases. (f) Phase-corrected SC aortic flow waveform following polynomial interpolation. (g) Schematic diagram of boundary condition specification.

## Reference

[1] Sakalihasan N, Michel J B, Katsargyris A, et al. Abdominal aortic aneurysms[J/OL]. Nature Reviews Disease Primers, 2018, 4(1): 34. DOI:10.1038/s41572-018-0030-7.

[2] Quintana R A, Taylor W R. Introduction to the Compendium on Aortic Aneurysms[J/OL]. Circulation Research, 2019, 124(4): 470–471. DOI:10.1161/CIRCRESAHA.119.314765.

[3] Kent K C. Abdominal Aortic Aneurysms[J/OL]. New England Journal of Medicine, 2014, 371(22): 2101–2108. DOI:10.1056/NEJMcp1401430.

[4] Miura S, Kurimoto Y, Maruyama R, et al. Initial two-day blood pressure management after endovascular aneurysm repair improves midterm outcomes by reducing the incidence of early type II endoleak[J/OL]. Journal of Vascular Surgery, 2024, 79(2): 251–259.e2. DOI:10.1016/j.jvs.2023.10.003.

[5] Nana P, Kouvelos G, Brotis A, et al. The effect of Endovascular Aneurysm Repair on Renal Function in Patients Treated for Abdominal Aortic Aneurysm[J/OL]. Current Pharmaceutical Design, 2019, 25(44): 4675–4685. DOI:10.2174/1381612825666191129094923.

[6] Varkevisser R R B, Swerdlow N J, de Guerre L E M V, et al. Midterm survival after endovascular repair of intact abdominal aortic aneurysms is improving over time[J/OL]. Journal of Vascular Surgery, 2020, 72(2): 556–565.e6. DOI:10.1016/j.jvs.2019.10.082.

[7] Derycke L, Avril S, Millon A. Patient-Specific Numerical Simulations of Endovascular Procedures in Complex Aortic Pathologies: Review and Clinical Perspectives[J/OL]. Journal of Clinical Medicine, 2023, 12(3): 766. DOI:10.3390/jcm12030766.

[8] Jayendiran R, Nour B, Ruimi A. Fluid-structure interaction (FSI) analysis of stent-graft for aortic endovascular aneurysm repair (EVAR): Material and structural considerations[J/OL]. Journal of the Mechanical Behavior of Biomedical Materials, 2018, 87: 95–110. DOI:10.1016/j.jmbbm.2018.07.020.

[9] Helo N, Chang A C, Hyun C, et al. Retrospective Review of Billowing Phenomenon—A Mimic of Endoleak Following Placement of Endologix Covered Stent for the Treatment of Abdominal Aortic Aneurysm[J/OL]. Annals of Vascular Surgery, 2017, 45: 239–246. DOI:10.1016/j.avsg.2017.06.127.

[10] Tran K, Feliciano K, Yang W, et al. Patient-specific changes in aortic hemodynamics is associated with thrombotic risk after fenestrated endovascular aneurysm repair with large diameter endografts[J/OL]. JVS: Vascular Science, 2022, 3. DOI:10.1016/j.jvssci.2022.04.002.

[11] Molony D S, Callanan A, Kavanagh E G, et al. Fluid-structure interaction of a patient-specific abdominal aortic aneurysm treated with an endovascular stent-graft[J/OL]. BioMedical Engineering OnLine, 2009, 8(1): 24. DOI:10.1186/1475-925X-8-24.

[12] Li Z, Kleinstreuer C. Blood flow and structure interactions in a stented abdominal aortic aneurysm model[J/OL]. Medical Engineering & Physics, 2005, 27(5): 369–382. DOI:10.1016/j.medengphy.2004.12.003.

[13] Li Z, Kleinstreuer C. Analysis of biomechanical factors affecting stent-graft migration in an abdominal aortic aneurysm model[J/OL]. Journal of Biomechanics, 2006, 39(12): 2264–2273. DOI:10.1016/j.jbiomech.2005.07.010.

[14] Zhang S, Laubrie J D, Mousavi S J, et al. 3D finite-element modeling of vascular adaptation after endovascular aneurysm repair[J/OL]. International Journal for Numerical Methods in Biomedical Engineering, 2022, 38(2): e3547. DOI:10.1002/cnm.3547.

[15] Laubrie J D, Mousavi J S, Avril S. A new finite-element shell model for arterial growth and remodeling after stent implantation[J/OL]. International Journal for Numerical Methods in Biomedical Engineering, 2020, 36(1): e3282. DOI:10.1002/cnm.3282.

[16] Peng C, He W, Huang X, et al. The study on the impact of AAA wall motion on the hemodynamics based on 4D CT image data[J/OL]. Frontiers in Bioengineering and Biotechnology, 2023, 11[2023-08-26]. https://www.frontiersin.org/articles/10.3389/fbioe.2023.1103905. DOI:10.3389/fbioe.2023.1103905.

[17] Arzani A, Wang J X, Sacks M S, et al. Machine Learning for Cardiovascular Biomechanics Modeling: Challenges and Beyond[J/OL]. Annals of Biomedical Engineering, 2022, 50(6): 615–627. DOI:10.1007/s10439-022-02967-4.

[18] Stalidzans E, Zanin M, Tieri P, et al. Mechanistic Modeling and Multiscale Applications for Precision Medicine: Theory and Practice[J/OL]. Network and Systems Medicine, 2020, 3(1): 36–56. DOI:10.1089/nsm.2020.0002.

[19] Talebi S, Madani M H, Madani A, et al. Machine learning for endoleak detection after endovascular aortic repair[J/OL]. Scientific Reports, 2020, 10(1): 18343. DOI:10.1038/s41598-020-74936-7.

[20] Yang Q, Hu J, Luo Y, et al. Detection of Endoleak after Endovascular Aortic Repair through Deep Learning Based on Non-contrast CT[J/OL]. CardioVascular and Interventional Radiology, 2024, 47(9): 1267–1275. DOI:10.1007/s00270-024-03805-x.

[21] Ayoub C, Appari L, Pereyra M, et al. Multimodal Fusion Artificial Intelligence Model to Predict Risk for MACE and Myocarditis in Cancer Patients Receiving Immune Checkpoint Inhibitor Therapy[J/OL]. JACC: Advances, 2025, 4(1): 101435. DOI:10.1016/j.jacadv.2024.101435.

[22] Li B, Feridooni T, Cuen-Ojeda C, et al. Machine learning in vascular surgery: a systematic review and critical appraisal[J/OL]. npj Digital Medicine, 2022, 5(1): 1–10. DOI:10.1038/s41746-021-00552-y.

[23] Wang Y R (Joyce), Yang K, Wen Y, et al. Screening and diagnosis of cardiovascular disease using artificial intelligence-enabled cardiac magnetic resonance imaging[J/OL]. Nature Medicine, 2024, 30(5): 1471–1480. DOI:10.1038/s41591-024-02971-2.

[24] Pezel T, Toupin S, Bousson V, et al. A Machine Learning Model Using Cardiac CT and MRI Data Predicts Cardiovascular Events in Obstructive Coronary Artery Disease[J/OL]. Radiology, 2025[2025-03-19]. https://pubs.rsna.org/doi/10.1148/radiol.233030. DOI:10.1148/radiol.233030.

[25] Kim S, Jiang Z, Zambrano B A, et al. Deep Learning on Multiphysical Features and Hemodynamic Modeling for Abdominal Aortic Aneurysm Growth Prediction[J/OL]. IEEE Transactions on Medical Imaging, 2023, 42(1): 196–208. DOI:10.1109/TMI.2022.3206142.

[26] Throop A, Bukac M, Zakerzadeh R. Prediction of wall stress and oxygen flow in patient-specific abdominal aortic aneurysms: the role of intraluminal thrombus[J/OL]. Biomechanics and Modeling in Mechanobiology, 2022, 21(6): 1761–1779. DOI:10.1007/s10237-022-01618-w.

[27] Caruso G, Martínez M Á, Peña E. Impact of mechanical properties of aneurysms and intraluminal thrombus on abdominal aortic aneurysm outcomes[J/OL]. Meccanica, 2025[2025-04-09]. 10.1007/s11012-025-01950-2. DOI:10.1007/s11012-025-01950-2.

[28] Bontekoe J, Matsumura J, Liu B. Thrombosis in the pathogenesis of abdominal aortic aneurysm[J/OL]. JVS-Vascular Science, 2023, 4: 100106. DOI:10.1016/j.jvssci.2023.100106.

[29] Wang D H J, Makaroun M S, Webster M W, et al. Effect of intraluminal thrombus on wall stress in patient-specific models of abdominal aortic aneurysm[J/OL]. Journal of Vascular Surgery, 2002, 36(3): 598–604. DOI:10.1067/mva.2002.126087.

[30] Thubrikar M J, Al-Soudi J, Robicsek F. Wall Stress Studies of Abdominal Aortic Aneurysm in a Clinical Model[J/OL]. Annals of Vascular Surgery, 2001, 15(3): 355–366. DOI:10.1007/s100160010080.

[31] van Griethuysen J J M, Fedorov A, Parmar C, et al. Computational Radiomics System to Decode the Radiographic Phenotype[J/OL]. Cancer Research, 2017, 77(21): e104–e107. DOI:10.1158/0008-5472.CAN-17-0339.

[32] Raut S S, Chandra S, Shum J, et al. The Role of Geometric and Biomechanical Factors in Abdominal Aortic Aneurysm Rupture Risk Assessment[J/OL]. Annals of Biomedical Engineering, 2013, 41(7): 1459–1477. DOI:10.1007/s10439-013-0786-6.

[33] Lindquist Liljeqvist M, Bogdanovic M, Siika A, et al. Geometric and biomechanical modeling aided by machine learning improves the prediction of growth and rupture of small abdominal aortic aneurysms[J/OL]. Scientific Reports, 2021, 11(1): 18040. DOI:10.1038/s41598-021-96512-3.

[34] Rezaeitaleshmahalleh S, Lyu Z, Mu N, et al. Characterization of small abdominal aortic aneurysms’ growth status using spatial pattern analysis of aneurismal hemodynamics[J/OL]. Scientific Reports, 2023, 13. DOI:10.1038/s41598-023-40139-z.

[35] Peng C, He W, Wang S, et al. Predict the endoleak risk after EVAR based on multi-dimensional anatomical features before AAA surgery[C/OL]//2024 IEEE International Conference on Cybernetics and Intelligent Systems (CIS) and IEEE International Conference on Robotics, Automation and Mechatronics (RAM). 2024: 435–440[2025-04-23]. https://ieeexplore.ieee.org/abstract/document/10673221. DOI:10.1109/CIS-RAM61939.2024.10673221.

[36] van Veldhuizen W A, Schuurmann R C L, IJpma F F A, et al. A Statistical Shape Model of the Morphological Variation of the Infrarenal Abdominal Aortic Aneurysm Neck[J/OL]. Journal of Clinical Medicine, 2022, 11(6): 1687. DOI:10.3390/jcm11061687.

[37] Bruse J L, Zuluaga M A, Khushnood A, et al. Detecting Clinically Meaningful Shape Clusters in Medical Image Data: Metrics Analysis for Hierarchical Clustering Applied to Healthy and Pathological Aortic Arches[J/OL]. IEEE Transactions on Biomedical Engineering, 2017, 64(10): 2373–2383. DOI:10.1109/TBME.2017.2655364.

[38] Geronzi L, Haigron P, Martinez A, et al. Assessment of shape-based features ability to predict the ascending aortic aneurysm growth[J/OL]. Frontiers in Physiology, 2023, 14: 1125931. DOI:10.3389/fphys.2023.1125931.

[39] Geronzi L, Martinez A, Rochette M, et al. Computer-aided shape features extraction and regression models for predicting the ascending aortic aneurysm growth rate[J/OL]. Computers in Biology and Medicine, 2023, 162: 107052. DOI:10.1016/j.compbiomed.2023.107052.

[40] Liu M, Dong H, Mazlout A, et al. The role of anatomic shape features in the prognosis of uncomplicated type B aortic dissection initially treated with optimal medical therapy[J/OL]. Computers in Biology and Medicine, 2024: 108041. DOI:10.1016/j.compbiomed.2024.108041.

[41] Scarpolini M A, Mazzoli M, Celi S. Enabling supra-aortic vessels inclusion in statistical shape models of the aorta: a novel non-rigid registration method[J/OL]. Frontiers in Physiology, 2023, 14[2023-11-09]. https://www.frontiersin.org/articles/10.3389/fphys.2023.1211461. DOI:10.3389/fphys.2023.1211461.

[42] Figueroa C A, Taylor C A, Yeh V, et al. Preliminary 3D computational analysis of the relationship between aortic displacement force and direction of endograft movement[J/OL]. Journal of Vascular Surgery, 2010, 51(6): 1488–1497. DOI:10.1016/j.jvs.2010.01.058.

[43] Horiguchi R, Takehara Y, Sugiyama M, et al. Postendovascular Aneurysmal Repair Increase in Local Energy Loss for Fusiform Abdominal Aortic Aneurysm: Assessments With 4D flow MRI[J/OL]. Journal of Magnetic Resonance Imaging, 2023, 57(4): 1199–1211. DOI:10.1002/jmri.28359.

[44] Zhang X, Mao B, Che Y, et al. Physics-informed neural networks (PINNs) for 4D hemodynamics prediction: An investigation of optimal framework based on vascular morphology[J/OL]. Computers in Biology and Medicine, 2023, 164: 107287. DOI:10.1016/j.compbiomed.2023.107287.

[45] Kang J, Li G, Che Y, et al. Four-dimensional hemodynamic prediction of abdominal aortic aneurysms following endovascular aneurysm repair combining physics-informed PointNet and quadratic residual networks[J/OL]. Physics of Fluids, 2024, 36. DOI:10.1063/5.0220173.

[46] Karthikesalingam A, Attallah O, Ma X, et al. An Artificial Neural Network Stratifies the Risks of Reintervention and Mortality after Endovascular Aneurysm Repair; a Retrospective Observational study[J/OL]. PLOS ONE, 2015, 10(7): e0129024. DOI:10.1371/journal.pone.0129024.

[47] Lee S R, Jabbour G, Schermerhorn M L. Increased Mortality After Endovascular Aortic Aneurysm Repair in Patients With Enlarged Aortic Necks[J/OL]. Journal of Vascular Surgery, 2024, 79(6): e125. DOI:10.1016/j.jvs.2024.03.156.

[48] Wang Y, Zhou M, Ding Y, et al. Development and Comparison of Multimodal Models for Preoperative Prediction of Outcomes After Endovascular Aneurysm Repair[J/OL]. Frontiers in Cardiovascular Medicine, 2022, 9[2025-03-19]. https://www.frontiersin.org/journals/cardiovascular-medicine/articles/10.3389/fcvm.2022.870132/full. DOI:10.3389/fcvm.2022.870132.

[49] Luo X, Wang J, Liang X, et al. Prediction of cerebral aneurysm rupture using a point cloud neural network[J/OL]. Journal of NeuroInterventional Surgery, 2023, 15(4): 380–386. DOI:10.1136/neurintsurg-2022-018655.

[50] Zhou M, Luo X, Wang X, et al. Deep Learning Prediction for Distal Aortic Remodeling After Thoracic Endovascular Aortic Repair in Stanford Type B Aortic Dissection[J/OL]. Journal of Endovascular Therapy: An Official Journal of the International Society of Endovascular Specialists, 2024, 31(5): 910–918. DOI:10.1177/15266028231160101.

[51] Peng C, Liu J, He W, et al. Numerical simulation in the abdominal aorta and the visceral arteries with or without stenosis based on 2D PCMRI[J/OL]. International Journal for Numerical Methods in Biomedical Engineering, 2022, 38(3): e3569. DOI:10.1002/cnm.3569.

[52] Raissi M, Yazdani A, Karniadakis G E. Hidden fluid mechanics: Learning velocity and pressure fields from flow visualizations[J/OL]. Science, 2020, 367(6481): 1026–1030. DOI:10.1126/science.aaw4741.

[53] Liang L, Mao W, Sun W. A feasibility study of deep learning for predicting hemodynamics of human thoracic aorta[J/OL]. Journal of Biomechanics, 2020, 99: 109544. DOI:10.1016/j.jbiomech.2019.109544.

[54] Jordanski M, Radovic M, Milosevic Z, et al. Machine Learning Approach for Predicting Wall Shear Distribution for Abdominal Aortic Aneurysm and Carotid Bifurcation Models[J/OL]. IEEE Journal of Biomedical and Health Informatics, 2018, 22(2): 537–544. DOI:10.1109/JBHI.2016.2639818.

[55] Yevtushenko P, Goubergrits L, Gundelwein L, et al. Deep Learning Based Centerline-Aggregated Aortic Hemodynamics: An Efficient Alternative to Numerical Modeling of Hemodynamics[J/OL]. IEEE Journal of Biomedical and Health Informatics, 2022, 26(4): 1815–1825. DOI:10.1109/JBHI.2021.3116764.

[56] Sarabian M, Babaee H, Laksari K. Physics-informed neural networks for brain hemodynamic predictions using medical imaging[J/OL]. DOI:10.1109/TMI.2022.3161653.

[57] Du P, Zhu X, Wang J X. Deep learning-based surrogate model for three-dimensional patient-specific computational fluid dynamics[J/OL]. Physics of Fluids, 2022, 34(8): 081906. DOI:10.1063/5.0101128.

[58] Pajaziti E, Montalt-Tordera J, Capelli C, et al. Shape-driven deep neural networks for fast acquisition of aortic 3D pressure and velocity flow fields[J]. PLOS COMPUTATIONAL BIOLOGY.

[59] Liang L, Liu M, Martin C, et al. A machine learning approach to investigate the relationship between shape features and numerically predicted risk of ascending aortic aneurysm[J/OL]. Biomechanics and Modeling in Mechanobiology, 2017, 16(5): 1519–1533. DOI:10.1007/s10237-017-0903-9.

[60] Williams J, Ozel A, Wolfram U. pyssam -- a Python library for statistical modelling of biomedical shape and appearance[A/OL]. arXiv, 2023[2023-09-08]. http://arxiv.org/abs/2301.04416.

[61] Lüdke D, Amiranashvili T, Ambellan F, et al. Landmark-free Statistical Shape Modeling via Neural Flow Deformations[A/OL]. arXiv, 2022[2023-09-14]. http://arxiv.org/abs/2209.06861.

[62] Bruce O L, Baggaley M, Welte L, et al. A statistical shape model of the tibia-fibula complex: sexual dimorphism and effects of age on reconstruction accuracy from anatomical landmarks[J/OL]. Computer Methods in Biomechanics and Biomedical Engineering, 2022, 25(8): 875–886. DOI:10.1080/10255842.2021.1985111.

[63] Battista F, Ficarelli R, Perrotta A, et al. The Fluid-Dynamics of Endo Vascular Aneurysm Sealing (EVAS) System failure[J/OL]. Cardiovascular Engineering and Technology, 2021, 12(3): 300–310. DOI:10.1007/s13239-021-00520-3.

[64] Sengupta S, Yuan X, Maga L, et al. Aortic haemodynamics and wall stress analysis following arch aneurysm repair using a single-branched endograft[J/OL]. Frontiers in Cardiovascular Medicine, 2023, 10[2025-04-21]. https://www.frontiersin.orghttps://www.frontiersin.org/journals/cardiovascular-medicine/articles/10.3389/fcvm.2023.1125110/full. DOI:10.3389/fcvm.2023.1125110.

